# Aperiodic parameters of the fMRI power spectrum associate with preterm birth and neonatal age

**DOI:** 10.1101/2024.12.16.24318785

**Authors:** Ilkka Suuronen, Silja Luotonen, Henry Railo, Antti Airola, Wajiha Bano, Harri Merisaari, Elmo P. Pulli, Isabella L.C Mariani Wigley, Elena Vartiainen, Niloofar Hashempour, Hasse Karlsson, Linnea Karlsson, Morten L. Kringelbach, Dafnis Batalle, Jetro J. Tuulari

**Author notes:** Correspondence to: Ilkka Suuronen, FinnBrain Birth Cohort study, Medisiina A3, Kiinamyllynkatu 4-8, 20520 Turku, Finland. These authors contributed equally to this work (Ilkka Suuronen, Silja Luotonen).

## Abstract

While strong associations of structural magnetic resonance imaging (sMRI) with preterm birth and post-menstrual age (PMA) have been reported, such associations for functional MRI (fMRI) have been considerably weaker. We studied the associations of the aperiodic parameters of neonatal fMRI Blood-oxygen-level-dependent (BOLD) signal power spectrum with preterm birth and PMA at scan using task-free fMRI data from the Developing Human Connectome Project (dHCP). First, we studied the associations of the aperiodic parameters of the BOLD signal from pre- and postcentral gyri with preterm birth and mapped the associations with PMA, postnatal age, and sex. Second, we used machine learning regression to predict PMA and postnatal age with 90 cortical and subcortical regions of interest (ROIs). We found clear differences between preterm and full-term groups, as well as males and females. Furthermore, our machine learning model predicted the age variables with relatively high accuracy (mean test R² = 0.20 – 0.41).

## Introduction

Brain development during gestation and after birth is rapid.^1^ The brain undergoes sequential, coordinated and hierarchical development that is also reflected in functional brain networks during the neonatal period and infancy.^2^ Previous work has demonstrated the presence and patterns of major brain networks functional connectivity in fetuses^3^ as well as in preterm^4–7^ and term infants.^8,9^ The maturation of motor and sensory systems occurs early^10^, and in infants, cortical functional networks exhibit large confinement to primary sensory and motor areas, suggesting a perception-action task-related functional network architecture.^10,11^

Preterm birth (birth before 37 weeks of gestation) is a common condition, occurring in about 1 in 10 births globally.^12^ It is associated with an increased likelihood for multiple adverse health outcomes in later life including brain, cognitive, and neural impairments.^4,13–15^ Considering brain maturation, preterm birth has been associated with altered brain development as younger gestational age at birth predicts smaller brain volumes^16–20^ and higher diffusivity in the prefrontal, parietal, motor, somatosensory, and visual cortices, likely suggesting delayed maturation of these cortical areas.^21^ Prior research has also demonstrated sex-specific alterations in preterm brain.^22^ In addition to the neural correlates of preterm birth, studies have unraveled relationships of many key steps of brain development with increasing postmenstrual age (PMA) at scan. However, the strength of the associations found in the earlier studies varies by brain metric. For instance, the associations between PMA and structural magnetic resonance imaging (MRI) metrics are quite strong (R^2^ ∼ 0.7)^23,24^ while functional MRI (fMRI) measures have shown much weaker associations (R^2^ ∼ 0.04).^25^ There is therefore a need for more accurate brain maturation assessment in neonates.

Electrophysiological activity of the brain exhibits both periodic (or oscillatory) and aperiodic (non-oscillatory) properties. The overall level of aperiodic activity, and how it changes as a function of frequency can be modelled with two parameters: offset and exponent, respectively.^26^ The parameters change as a function of age, with the exponent decreasing in infants^27^, and both exponent and offset decreasing during the early-childhood^28,29^ and early-adulthood.^30^ Based on these EEG studies, we propose that the parameters of aperiodic brain activity could serve as markers for the pace of (functional) brain maturation. Given that aperiodic brain activity is assumed to be ‘scale-free’ (i.e., it is not restricted to a specific temporal scale)^31^, age-related changes could also be detected using aperiodic parameters of the resting-state functional MRI (rs-fMRI) blood-oxygen-level-dependent (BOLD) signal. While the utility of parameterizing brain activity in terms of its periodic and aperiodic components has been demonstrated in numerous electrophysiological studies^26,27,32,33^, the methodology has not been widely adopted for the analysis of fMRI datasets.

In the current study, we parameterized the aperiodic components of the rs-fMRI BOLD signal power spectra, filtered to a narrowband of 0.01–0.15 Hz, from 90 cortical and subcortical regions of interest (ROIs) in terms of their exponent and offset. We then mapped the associations between the parameters pertaining to the pre- and postcentral gyri and preterm birth, and postmenstrual and postnatal age as well as sex were included to linear mixed effect models. Finally, we performed data driven machine learning based regression to predict postmenstrual and postnatal age from the parameters of all ROIs. Based on previous electrophysiological studies^27–29,34^ and diffusion tensor imaging studies in preterm infants^21^, we hypothesized that the postmenstrual and postnatal ages of neonates are negatively associated with aperiodic parameters and that preterm born neonates have higher aperiodic parameters compared to term-born neonates, reflecting a delayed neural developmental processes in preterm born neonates.

## Materials and methods

The current study used preprocessed data released as part of the Developing Human Connectome Project (dHCP) data release 3.0, and the methods are described in detail previously.^25^

Research participants were prospectively recruited as part of the dHCP, an observational, cross-sectional Open Science programme approved by the UK National Research Ethics Authority (14/LO/1169). Written consent was obtained from all participating families prior to imaging. Term-born neonates were recruited from the postnatal wards and approached on the basis of being clinically well. Preterm-born neonates were recruited from the neonatal unit and postnatal wards. Neonates were not approached for study inclusion if there was a history of severe compromise at birth requiring prolonged resuscitation, a diagnosed chromosomal abnormality, or any contraindication to MRI scanning (e.g., due to incompatible implants). No neonates included in the final study group required treatment for a clinically significant brain injury. The latest data release includes a total of 887 datasets from 783 neonates including healthy term-born neonates, preterm neonates and neonates at high risk for atypical neurocognitive development.^35^

### Participants

The openly available dHCP data set (third release) consists of 887 rs-fMRI scans from 783 neonatal research subjects between 26-45 weeks of post-menstrual age (359 females), 205 of whom were born preterm. Minor incidental findings were not considered an exclusion criterium. For a more detailed description of the data set, see Edwards et al.^35^ 105 scans were flagged for failed fMRI quality control and excluded from the current analyses. Further, 129 scans were from non-singleton participants, and excluded from current analyses to not cause indirect information leakage. Finally, we only included the last non-flagged scan from each participant, resulting in 605 unique, non-flagged scans from singleton participants. The FOOOF model fit failed for six participants, for which reason the analyses were performed on a subset of 599 participants (276 female, PMA 27–45 weeks), 116 of them preterm-born. For a more detailed characterization of the subset used in analyses, see Table 1.

**Table 1.**
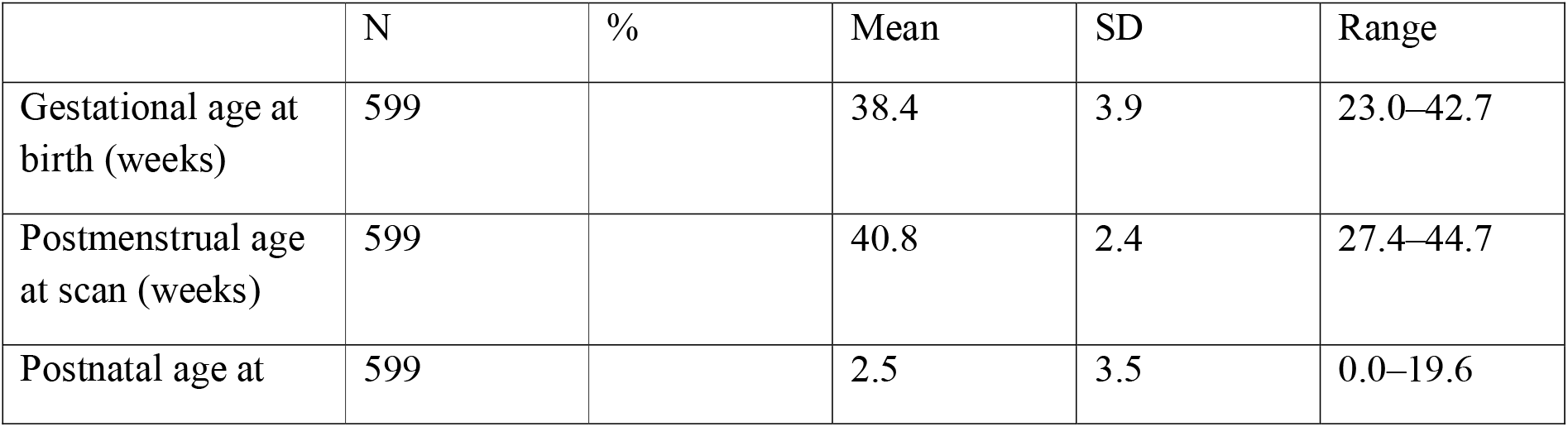
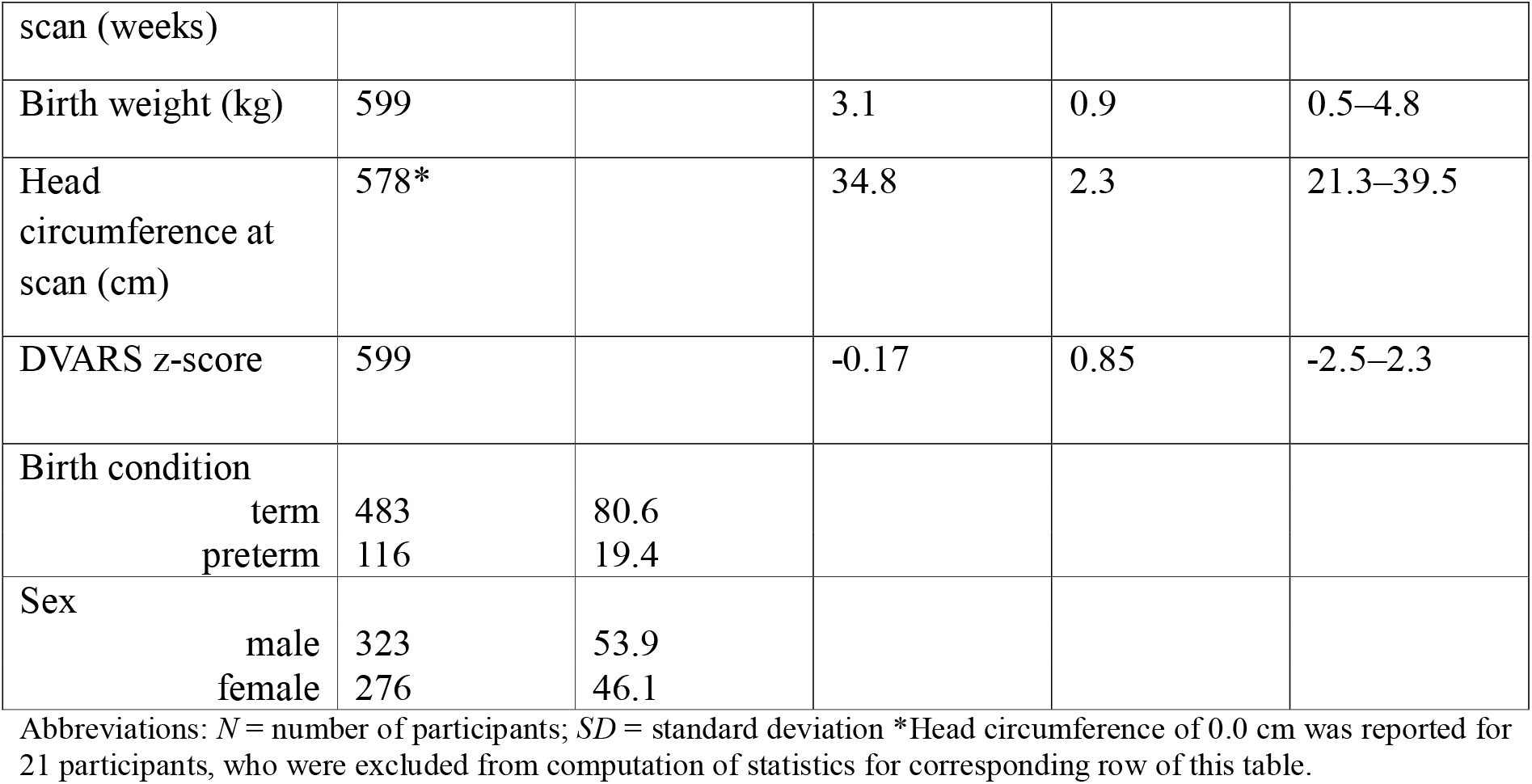
Characteristics of participants included in the analyses.

### fMRI data acquisition and preprocessing

The MR imaging was carried out using a 3T Philips Achieva scanner running modified Release 3.2.2 software. The length of the fMRI acquisition was 15 minutes, and was collected with parameters: TE/TR = 38/392 ms, 2300 volumes, with an acquired spatial resolution of 2.15 mm isotropic.^35^ Participants were scanned non-sedated using Neonatal Brain Imaging System (NBIS), consisting of a dedicated 32-channel array coil and a positioning device.^36^ The functional data imaging acquisition used optimized multiband sequence tuned for neonatal participants^37^ with phase optimized multiband pulses used throughout.^38^ For detailed descriptions of the automated processing pipeline and the motion and distortion correction techniques applied to all dHCP open access pre-processed fMRI data, see Fitzgibbon et al.^39^ and Andersson et al.^40–43^

### Derived brain measures

An algorithmic method known as FOOOF (Fitting Oscillations and One-Over-F) for parameterizing the power spectrum into periodic and aperiodic component parameters has been suggested.^26^ While the periodic component is characterized by such parameters as its center frequency, power and bandwidth, the 1/f-like aperiodic component is characterized by its offset and exponent parameters. The performance of the algorithm in characterizing neural power spectra has been validated against both simulated power spectra with known ground truth and human expert labeling on real EEG and simulated data, demonstrating low error rate on both test arrangements.^26^

The FOOOF algorithm parameterizes the power spectrum into its periodic and aperiodic components by creating an initial fit of the aperiodic component and subtracting it from the total power spectrum resulting in a flat spectrum, from which in an iterative process maxima are identified as peaks and used for fitting gaussians which are then subtracted from the flattened spectrum. When no further peaks surpassing a threshold value can be identified, a final periodic multi-gaussian model is fit using all maxima thus identified. The periodic model is subtracted from the original spectrum, resulting in a noisy aperiodic component, to which the final aperiodic model is fit. The aperiodic model specifically is characterized by offset and exponent parameters as per the following formula:

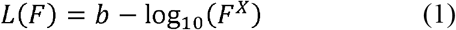

Where F refers to frequency, b to the offset parameter and X to the exponent variable. Notably, power values are presented in a logarithmic scale, for which reason negative offset values are possible.

For each time series representing a ROI specific BOLD signal, we estimated and parameterized the power spectrum between 0.01 and 0.15 Hz using multitaper^44^ spectral analysis, and the FOOOF algorithm (Python implementation version 1.1.0), resulting in parameters for the periodic and aperiodic components of the signal, as well an R² score for the goodness of fit for each ROI. For a summary on frequency range selection in earlier studies on BOLD signal, see e.g., Glerean et al.^45^ As the frequency range of interest was so narrow, we set the minimum width of oscillation peaks to 0.001 Hz while specifying no maximum width. Based on earlier work studying the characteristics of the BOLD signal power spectrum, we set the threshold value for oscillatory peaks to 1.5 SD over the mean, as well as the maximum number of peaks to find to two.^46^

### Linear mixed effect regression models

Statistical analyses were performed using RStudio (2022.07.1+554) and JASP (2022, version 0.16.3). To estimate the associations between the aperiodic parameters (exponent or offset) of fMRI BOLD signal and preterm birth, sex, PMA at scan and postnatal age, the *lmer* function from *afex* package of RStudio was used. Linear mixed effects regression model analyses were performed using maximum likelihood estimation. Fixed effects were PMA at scan, postnatal age, sex, and premature birth as well as hemisphere (right/left). Interaction terms with ROI (precentral gyri / postcentral gyri) were included. In addition, in the offset model (Model 2) exponent was included as a fixed effect to control the possible effects of exponent changes on offset values. The intercepts of subjects were modeled as random effects, including random slopes for the motion outliers. The linear mixed effects regression models had the following structure:

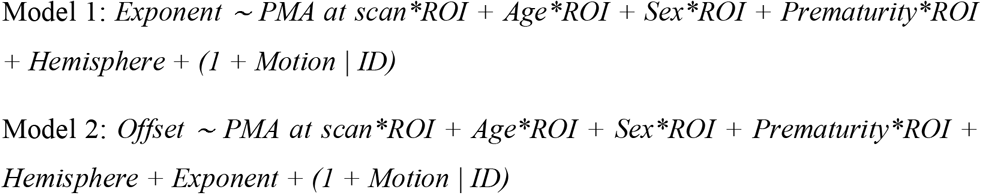

PMA at scan means the postmenstrual age (PMA) of the neonate at scan (weeks; z-transformed) and Age means the postnatal age of the child (weeks; z-transformed). Sex means the sex of the child (male = 0, female = 1) and Prematurity means term versus preterm birth (term-born = 0, preterm = 1; postmenstrual age at birth < 37 weeks). ROI means region-of-interest, either postcentral (= 0) or precentral (= 1) gyri and Hemisphere means left (= 0) or right (= 1) side of the brain. Motion means the number of motion-compromised volumes (outliers based on DVARS; z-transformed). In Model 2, the exponent was z-transformed.

The models were originally run with only main effects of independent variables included, but as interaction terms with ROI were added into the models, the fit of the models improved (based on lower Akaike information criterion (AIC) and statistically significant result of likelihood ratio test before vs. after interaction term inclusion). The Prematurity variable had moderate correlation with other age variables (for correlation matrix of all variables included in models: see Figure S1 in Supplemental file 1), but lower AIC (when Prematurity was included) supported the inclusion of the Prematurity as a predictor. Likelihood ratio test (Prematurity not included vs. included) was not statistically significant in Model 1 (p = 0.09) and in Model 2 (p = 0.06). Possibility of singularity of the models were checked, and not detected.

Further, the model assumptions were visually assessed for both models. At first, the Q-Q plot of the Model 1 showed slightly S-shaped pattern, while other visual assessments supported normal distribution of the residuals. After residual outlier detection and exclusion (± 3SD, N = 26), the Q-Q plot improved. In Model 2, visual assessments supported normal distribution of the residuals and the need for outlier exclusion was not established. For more information about model diagnostics, please see Figures S2-S5 and Tables S1-S2 in Supplemental file 1.

Finally, to plot the association between PMA and exponent / offset, quadratic term for PMA (PMA^2^) were added to Model 1 and 2 (see “Quadratic Mixed Effect Models” in Supplemental file 1).

### Linear regression models

To map the estimates of *Prematurity* (term-born = 0, preterm = 1) for aperiodic parameters (exponent or offset) of fMRI BOLD signal across 90 different cortical and subcortical brain areas, the linear models following the formulas of the linear mixed effect regression models described above were used. For more detailed description, see “Linear Regression Models” in Supplemental file 1.

### Machine learning models

The machine learning based data-analysis was performed using Python 3.8.3^47^ in conjunction with external libraries Numpy 1.23.5^48^, Pandas 2.0.3^49^, Neurokit2 0.2.7^50^ and Scikit-learn 1.2.2.^51^ The script used for performing the machine learning based regression analyses is available in https://gitlab.utu.fi/ilksuu/fmri_aperiodic_parameters_neonatal_age_prediction.

We predicted the participants’ PMA and postnatal age as target variables using a supervised machine learning regression model. For each target variable, we trained and tested a regression model with either exponent, offset or both kinds of parameters pertaining to all ROIs as predictors using repeated and nested 10-fold cross-validation algorithm. To control for the effect of preterm birth, we trained and tested the regression models separately for the full sample and term-born only participants. In addition to the exponent and offset parameters of the aperiodic component of the power spectra of the BOLD signals, no additional predictors were introduced to the machine learning models. Performing the machine learning based regression analysis separately using three sets of predictors (i.e., offset, exponent and both) on two subsets (term-born only and all participants) of the data in predicting two different age target variables (postnatal and postmenstrual age at scan) resulted in 12 predictive models in total.

We used ElasticNet^52^, an l1- and l2-regularized linear regression model to make predictions on the target variables and rank the predictors for interpretation using the models’ beta coefficients. While l1-regularized regression (aka. LASSO) performs feature selection by enforcing sparsity, thus making the model more interpretable, it can lead to model instability if the features are highly correlated, which typically is the case with neuroimaging data.^53^ In contrast, ElasticNet accomplishes a grouping effect via l2-regularization, which is known to correct the instability-inducing tendency for arbitrary choice between correlated predictors^54^ and is considered a standard predictive algorithm in machine learning based neuroimaging studies.^53^ ElasticNet finds the optimal model coefficients by minimizing β in the following formula:

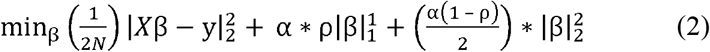

Where β stands for the beta coefficients or “weights” of the model, α stands for the overall regularization strength and ρ stands for the ratio of l1-norm used in the regularization.

To estimate the models’ predictive performance on previously unseen data with low bias and variance, we utilized 10-times repeated non-stratified nested k-fold cross-validation algorithm, with 10 outer and 5 inner folds for the purpose of grid search -based hyperparameter tuning, the hyperparameters in question being the “α” and “ρ” in the ElasticNet formula (2) (i.e., “alpha” and “l1_ratio” respectively, in the scikit-learn implementation). The beta coefficients of the trained models were averaged over folds and ranked based on average coefficient value for the purpose of feature importance analysis. For a discussion on the rationale for using nested cross-validation for unbiased model evaluation, see, e.g., Cawley and Talbot.^55^

Prior to fitting the ElasticNet model, the data was standardized by subtracting the mean and scaling to unit norm – a recommended preprocessing step that is considered to have the effect of increasing the interpretability of the model.^53^ This preprocessing operation was performed in runtime and separately for each cross-validation fold in order to avoid data leakage from test set to the model, which could occur if such preprocessing was performed prior to splitting the data.

### Data availability

The data that support the findings of this study are openly available in The National Institute of Mental Health Data Archive at https://nda.nih.gov/edit_collection.html?id=3955, reference number 3955.

## Results

The aperiodic fit was generally acceptable across the ROIs, but it was highest for somatosensory areas (Figure 1). There was also a trend of poorer fits towards the inferior parts of the brain that may be due to poor signal to noise ratio in those regions.^39^

**Figure 1.**
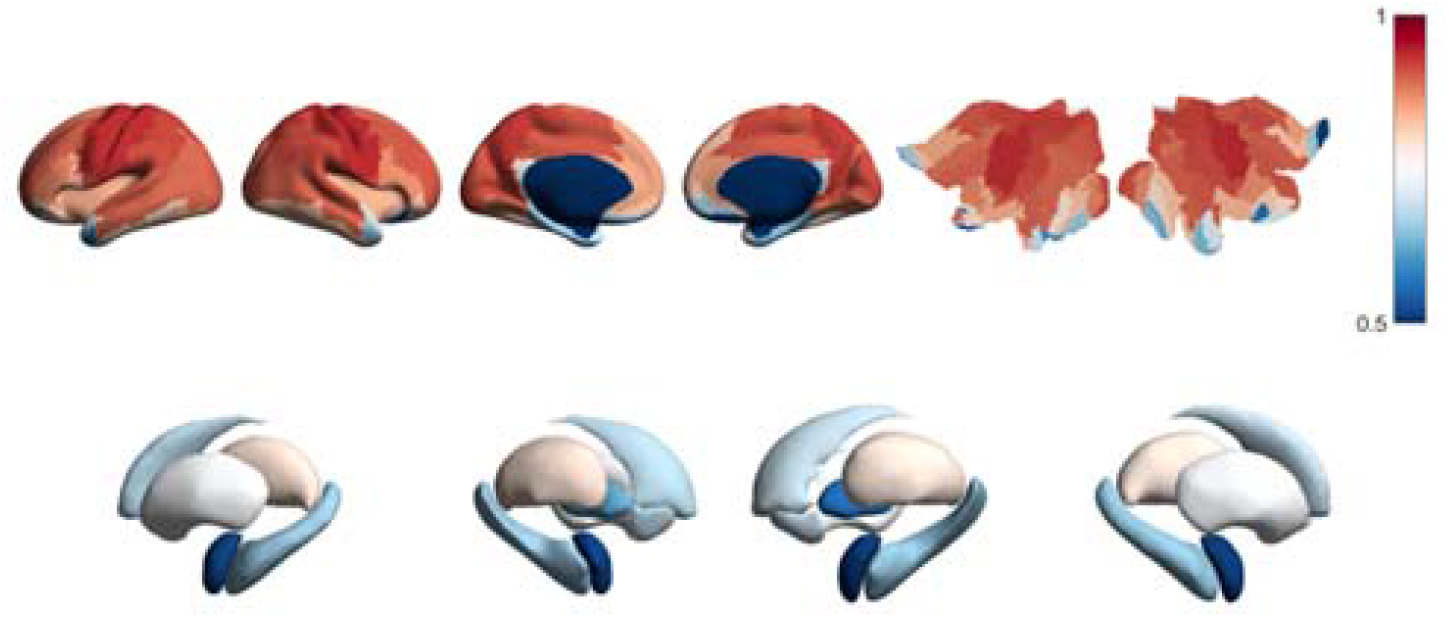
The FOOOF model fit group mean R^2^ values across the AAL areas.

### Effects of postmenstrual age at scan

Postmenstrual age at scan was positively associated with the offset (*estimate* 0.06, *SE* 0.01, *p* < 0.001). Further, interaction effects showed that this association was statistically significantly dependent on ROI ((Table 1B: *estimate* 0.04, *SE* 0.01, *p* < 0.001). This means that the increase of one standard deviation (*SD*) in child PMA predicted a +0.06 change in offset in postcentral gyri and a +0.10 change in precentral gyri. In quadratic model, association between PMA and offset was nearly linear as presented in Figure S1 in Supplemental file 3.

The main effect of PMA at scan was not statistically significantly associated with the exponent, but its interaction effect with ROI was statistically significant (Table 1A: *estimate* –0.03, *SE* 0.01, *p* = 0.002). This means that there was a crossover interaction, i.e., the association between PMA and exponent was dependent on the ROI. In postcentral gyri, the increase of one SD in child PMA predicted +0.02 change in exponent, while in precentral gyri, the corresponding change was negligible, –0.01. To see the relationship between predicted aperiodic parameters and child PMA, see Figure S2 in Supplemental file 3. In quadratic model, association between PMA and exponent was reverse U-shaped as presented in Figure S3 in Supplemental file 3.

### Effects of postnatal age

The negative association between child’s postnatal age and exponent was statistically significant (Table 2A: *estimate* –0.17, *SE* 0.03, *p* < 0.001). This association did not differ statistically significantly between the pre- and postcentral gyri, meaning that the increase of one *SD* in postnatal age predicts –0.17 change in exponent in both ROIs.

**Table 2.** Results of the linear mixed-effects regression model for exponent (Table 2A) and Offset (Table 2B).

| **Table 2A** | Estimate | SE | df | t value | p value |
| --- | --- | --- | --- | --- | --- |
| Intercept | 1.89 | 0.03 | 640 | 69.59 | < 0.001 |
| PMA at scan | 0.02 | 0.02 | 611 | 0.87 | 0.385 |
| ROI | 0.01 | 0.01 | 1771 | 1.65 | 0.099 |
| Postnatal age | -0.17 | 0.03 | 611 | -6.04 | < 0.001 |
| Sex | -0.09 | 0.03 | 632 | -2.76 | 0.006 |
| Prematurity | -0.17 | 0.08 | 631 | -2.11 | 0.036 |
| Hemisphere | 0.01 | 0.01 | 1770 | 1.66 | 0.098 |
| PMA at scan : ROI | -0.03 | 0.01 | 1776 | -3.12 | 0.002 |
| Postnatal age : ROI | -0.01 | 0.01 | 1772 | -1.26 | 0.208 |
| Sex : ROI | 0.02 | 0.01 | 1770 | 1.44 | 0.150 |
| Prematurity : ROI | 0.03 | 0.03 | 1771 | 1.05 | 0.295 |
| **Table 2B** | Estimate | SE | df | t value | p value |
| Intercept | -3.50 | 0.02 | 749 | -209.47 | < 0.001 |
| PMA at scan | 0.06 | 0.01 | 653 | 4.07 | < 0.001 |
| ROI | 0.12 | 0.01 | 1795 | 12.16 | < 0.001 |
| Postnatal age | -0.02 | 0.02 | 664 | -1.42 | 0.156 |
| Sex | 0.04 | 0.02 | 700 | 2.08 | 0.038 |
| Prematurity | -0.09 | 0.05 | 686 | -1.91 | 0.057 |
| Hemisphere | 0.01 | 0.01 | 1794 | 1.98 | 0.048 |
| PMA at scan : ROI | 0.04 | 0.01 | 1799 | 4.88 | < 0.001 |
| Postnatal age : ROI | -0.02 | 0.01 | 1794 | -1.98 | 0.048 |
| Sex : ROI | 0.02 | 0.01 | 1793 | 1.87 | 0.062 |
| Prematurity : ROI | 0.06 | 0.03 | 1794 | 1.98 | 0.048 |

The negative effect of postnatal age on offset was not statistically significant. However, its interaction effect with ROI was statistically significant (Table 2B: *estimate* –0.02, *SE* 0.01, *p* = 0.048). The result indicates that an increase of one standard deviation in postnatal age decreases the difference in offset values between the precentral and postcentral gyri by –0.02, resulting in the region-related difference disappearing at approximately 20 weeks of postnatal age. To see the relationship between predicted aperiodic parameters and child postnatal age, see Figure S1 in Supplemental file 3.

### Effects of prematurity

Prematurity was negatively associated with the exponent (Table 2A: *estimate* = –0.17, *SE* 0.08, *p* = 0.036). This means that exponent values were statistically significantly smaller (– 0.17 difference) in neonates born preterm compared to term-born neonates.

Prematurity was nearly statistical significantly associated to offset (Table 2B: *estimate* −0.09, *SE* 0.05, *p* = 0.057). If true, this would mean that offset values are in general 0.09 smaller in preterm neonates compared to term born neonates. However, the interaction effect of prematurity and ROI was statistically significant (Table 2B: *estimate* 0.06, *SE* 0.03, *p* = 0.048), indicating that effect of prematurity is even weaker in precentral gyri. To see the differences in predicted aperiodic parameters between the term-born versus preterm neonates visualized, see Figure 2.

**Figure 2.**
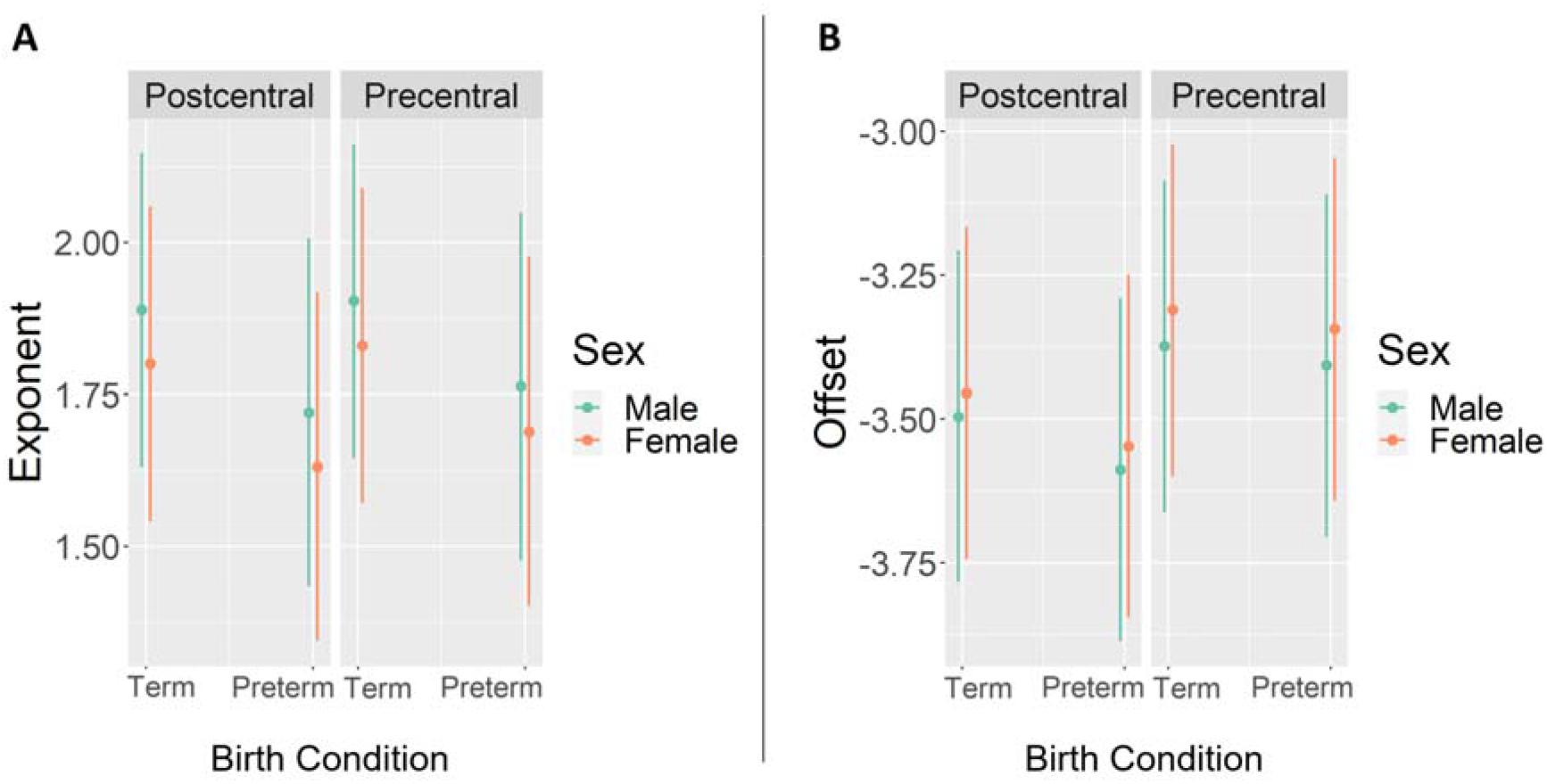
Predicted aperiodic parameters (conditioned on random effects) in term-born vs. preterm neonates and females versus males from the pre- and postcentral gyri. (**A**) Aperiodic exponents. (**B**) Aperiodic offsets.

To see the cortical and subcortical projected estimates of prematurity from the linear regression models (exponent / offset) across all 90 different brain areas, please see the Figure 3. After FDR correction, effect of prematurity on aperiodic exponent remained statistically significant only in two brain areas: right hippocampal (*adjusted p* = 0.009) and right parahippocampal (*adjusted p* = 0.029) areas. No statistically significant effects of prematurity on aperiodic offset remained in any of the brain areas. All estimates, their p values, and FDR corrected p values are presented in Table S1 and S2 in Supplemental file 3.

**Figure 3.**
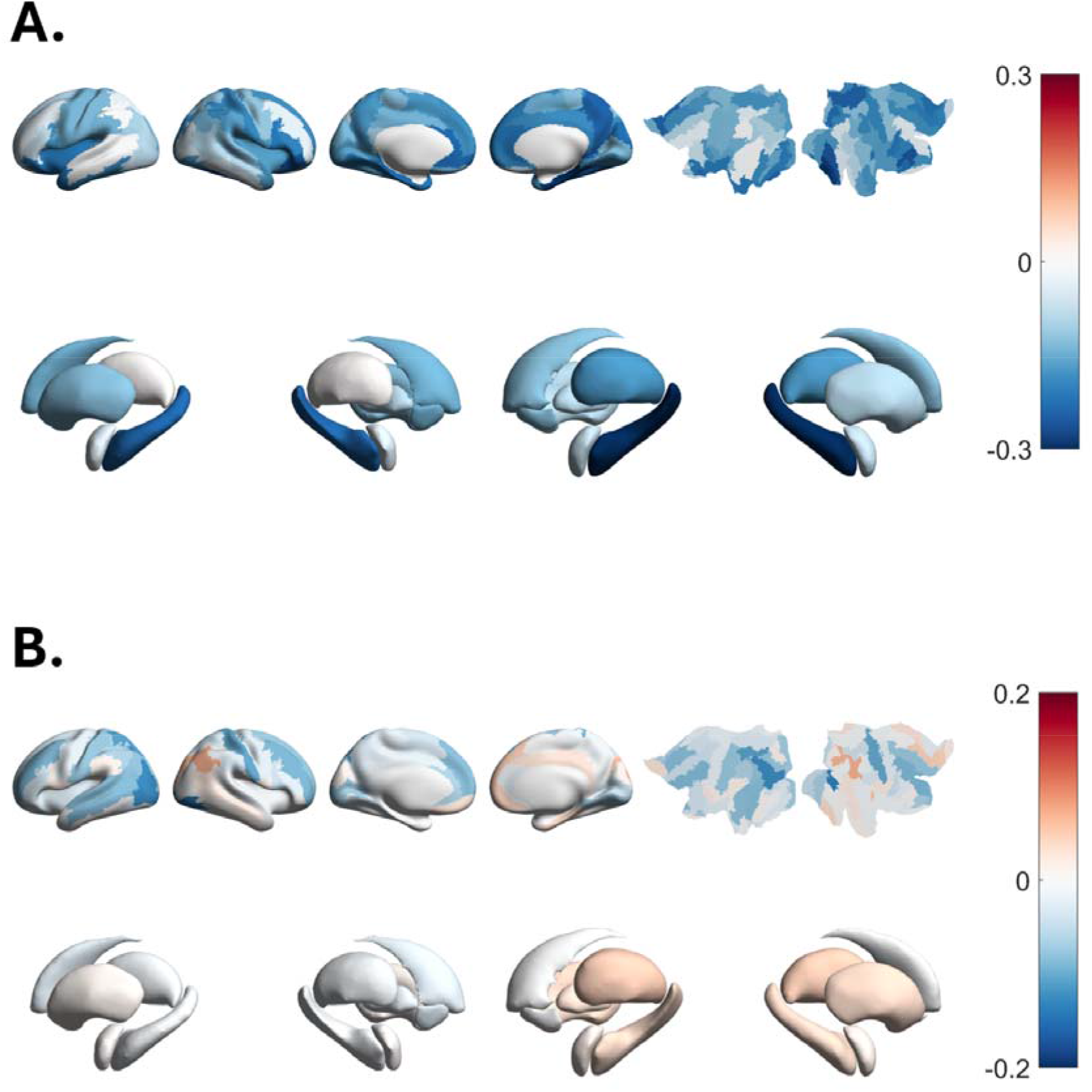
Estimates from linear models for preterm birth (0 = term-born, 1 = preterm), projected onto cortex and subcortex. (**A**) Aperiodic exponent. (**B**) Aperiodic offset.

### Effects of sex

In females, aperiodic exponents were smaller (Table 2A: *estimate* = –0.09, *SE* 0.03, *p* = 0.006) and aperiodic offsets higher compared to males (Table 2B*: estimate* = 0.04, *SE* 0.02, *p* = 0.038). No statistically significant interactions between child’s sex and ROI were found.

### Regional and hemisphere-specific differences

The offset values were higher in precentral gyri when compared to postcentral gyri (Table 2B: *estimate* 0.12, *SE* 0.01, *p* < 0.001). There was no statistically significant association between ROI (postcentral / precentral gyri) and exponent. In the right hemisphere, offset values were negligibly 0.01 higher than in left hemisphere (Table 2B: *estimate* = 0.01, *SE* = 0.01, *p* = 0.048). Exponent values were not associated with the side of the hemisphere (right/left).

### Machine learning results

Predictive model performance was estimated separately using only offset-based predictors, only exponent-based predictors, and both types of predictors at once. Performance was also estimated separately for postnatal age and PMA at the time of scan as target variables, as well as for subset of the data containing only the term-born participants and all participants to control for the effect of preterm birth. For an estimate of the model’s generalization ability, the mean values of the repeatedly cross-validated test and train set performance estimates over the cross-validation folds were reported in terms of the coefficient of determination (*R²*) and the mean absolute error (*MAE*) in weeks of age. For the performance estimates for each of the different analysis settings, see Table 3 A-C.

**Table 3.** Cross-validated performance estimates. Reported in terms of mean *R²* and *MAE* (in weeks). N = 599 neonates (483 term-born).

| **Table 3A** Cross-validated performance estimates for analyses predicting postnatal and postmenstrual age at scan using offset features. |  |  |  |  |
| --- | --- | --- | --- | --- |
|  | term-born postnatal | full sample postnatal | term-born postmenstrual | full sample postmenstrual |
| Train $R^2$ | 0.502 | 0.309 | 0.475 | 0.502 |
| Test $R^2$ | 0.363 | 0.238 | 0.320 | 0.350 |
| Train MAE | 0.740 | 1.828 | 0.955 | 1.286 |
| Test MAE | 0.826 | 1.884 | 1.075 | 1.420 |
| **Table 3B** Cross-validated performance estimates for analyses predicting postnatal and postmenstrual age at scan using exponent features. |  |  |  |  |
|  | term-born postnatal | full sample postnatal | term-born postmenstrual | full sample postmenstrual |
| Train $R^2$ | 0.483 | 0.299 | 0.436 | 0.393 |
| Test $R^2$ | 0.358 | 0.224 | 0.307 | 0.204 |
| Train MAE | 0.749 | 1.822 | 0.983 | 1.369 |
| Test MAE | 0.821 | 1.886 | 1.077 | 1.513 |
| **Table 3C** Cross-validated performance estimates for analyses predicting postnatal and postmenstrual age at scan using both offset and exponent features. |  |  |  |  |
|  | term-born postnatal | full sample postnatal | term-born postmenstrual | full sample postmenstrual |
| Train $R^2$ | 0.579 | 0.318 | 0.565 | 0.668 |
| Test $R^2$ | 0.403 | 0.243 | 0.354 | 0.411 |
| Train MAE | 0.673 | 1.805 | 0.864 | 1.043 |
| Test MAE | 0.789 | 1.868 | 1.039 | 1.319 |

The cross-validated mean performance scores associated with the different settings of input and output variables demonstrate moderate to relatively high test performance for all such settings (Test *R²’s* 0.20–0.41). Offset predictors yielded generally somewhat higher performance estimates (0.24–0.36) for predicting both types of target variable than exponent predictors (0.20–0.36). Unsurprisingly the models trained with both types of predictors at once performed better than either type separately (0.24–0.41). As for the target variables, the models predicting postnatal age seem to perform better on term-born only subsample (0.36– 0.40) as compared to full sample (0.22–0.24), whereas models predicting PMA seem to perform better on full sample (0.20–0.41) than term-only subsample (0.31–0.35), with the exception of exponent predictor model predicting PMA (0.20). Comparing mean train performance scores to test performance scores demonstrates some amount of model overfitting despite the regularization mechanism inherent to the ElasticNet.

For the purpose of model interpretation via feature importance analysis, Figures S2 and S3 in Supplemental file 2 display the top-ranking positive and negative mean ElasticNet coefficients, while Figure 4 displays all mean ElasticNet coefficients projected onto corresponding regions of interest on the cortex and subcortex, across all cross-validation folds for each of the analysis settings on full-term subjects. For complete reports on ElasticNet mean coefficient values for each analysis setting, see Figures S4-11 in Supplemental file 2.

**Figure 4.**
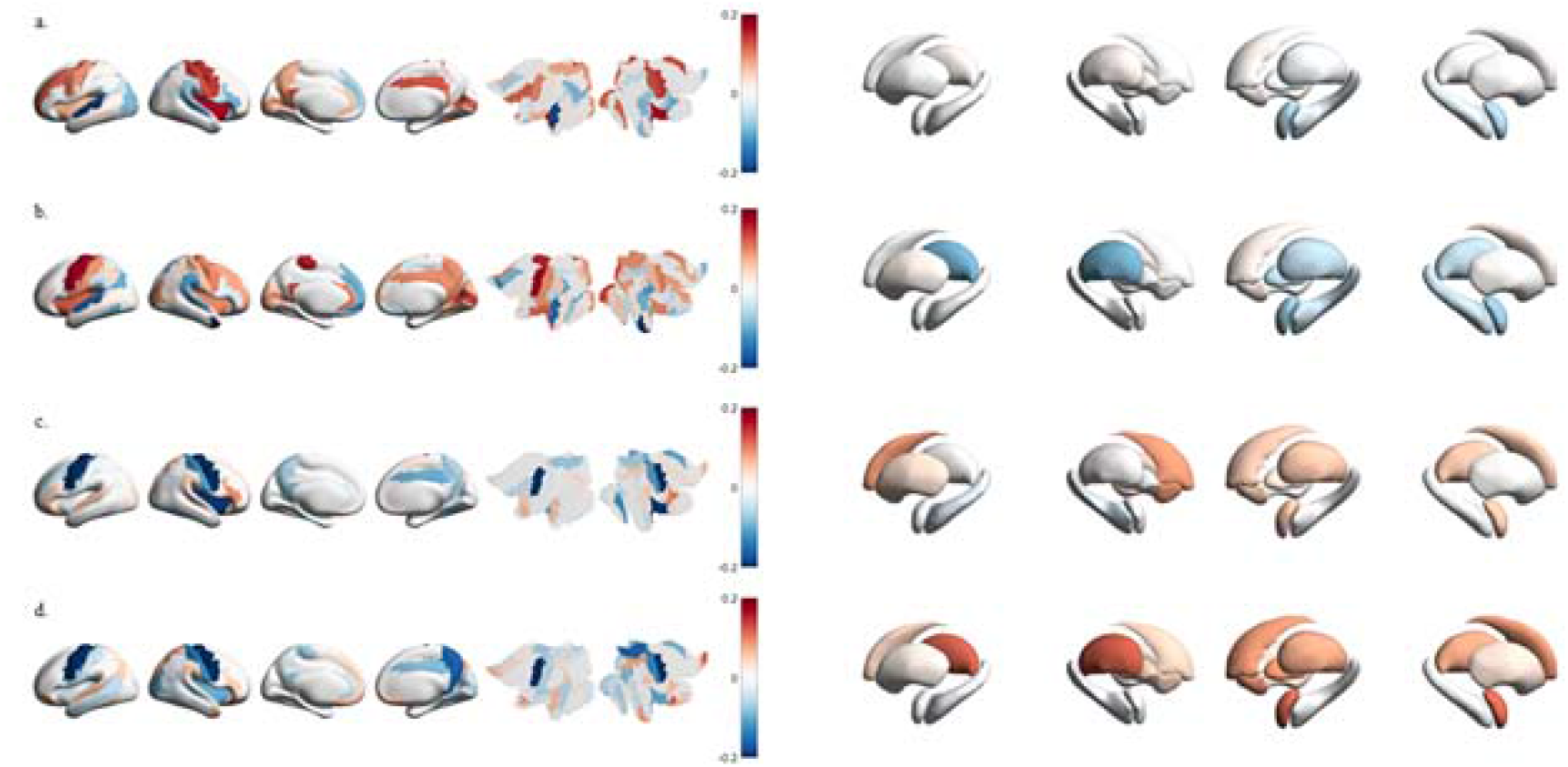
Model beta coefficients for predicting the age of term-born participants on different analysis settings, projected onto cortex and subcortex. (**A**) predicting postnatal age with offsets. (**B**) Predicting postmenstrual age with offsets. (**C**) Predicting postnatal age with exponents predictors. (**D**) Predicting postmenstrual age with exponents.

## Discussion

In this cross-sectional study, we used the aperiodic parameters of the BOLD signal power spectrum as a prospective index of brain maturation. Prematurity associated with lower offsets and exponents of aperiodic activity compared to term-born neonates. This reflects a flatter distribution of aperiodic signal activity (i.e., flattening power spectrum) across frequencies. The difference between preterm vs. term-born neonates was narrowly non-significant in offsets *(p* = 0.057) and significant in exponents (*p* = 0.036). The offsets were positively associated with PMA and negatively with postnatal age while for the exponents the association with PMA was not statistically significant and the association to postnatal age was strongly negative. Our findings indicate an increase in aperiodic activity with postmenstrual aging, followed by a decrease in the postnatal period (as presented in Figures S1 & S3 in Supplemental file 3). Finally, in our data driven machine learning based analyses, we observed relatively strong associations (*R²’s* 0.20–0.41) between aperiodic parameters of rs-fMRI BOLD signal and neonate age, demonstrating the feasibility of the methodology for studying functional brain maturation.

### Postmenstrual age and aperiodic parameters

During gestation, there is a proliferation of neuroblasts and synaptogenesis, ultimately leading to the capability of neurons to fire repetitive action potentials. Additionally, spontaneous neural activity begins to occur in mid-gestation (approximately at gestation week 18).^56,57^ Given that asynchronous signals (i.e., offset) have been identified as a principal source of BOLD responses^58^ and further, electrophysiological power shifts have been shown to be reliable predictor of neuronal spikes^59^, our results could reflect increasing neuronal firing rates as a function of postmenstrual age during gestation.

We chose the somatosensory regions for closer examination as they are known to be among the regions that mature early.^3^ Offset values were higher in precentral compared to postcentral gyri, indicating greater power of the BOLD signal in the precentral gyri. Considering offset as an indirect proxy of neural activity, this suggests higher neural activity in the precentral (motor) gyri compared to postcentral (somatosensory) gyri. However, this difference narrows down after birth as postnatal age increases. We observed an increase in exponent values in the postcentral gyri as a function of PMA while in precentral gyri, no noticeable change was detected. This suggests that the aperiodic activity distribution steepens in the postcentral gyri as a function of postmenstrual age. Given that in previous EEG studies, flattening of the power spectrum has been linked to postnatal aging^27–30,34^, our results could indicate earlier maturation of the precentral (motor) gyri compared to postcentral (somatosensory) gyri during gestation. This aligns with an earlier diffusion tensor imaging study of preterm newborns, which found higher fractional anisotropy and lower diffusivity in motor tracts compared to sensory tracts, reflecting earlier white matter maturation of motor system compared to sensory pathways.^60^

It is important to note that our study focused on changes in the low-frequency power spectrum of the BOLD signal (0.01–0.15 Hz), and caution is warranted when comparing our results to prior EEG literature. However, previous studies on fractional low-frequency fluctuation (fALFF) activation in adults have reported negative correlations between age and the fALFF activation in adults^61,62^, and fALFF activation has been demonstrated to be a valid predictor of age in children.^63^ These findings suggest a trend towards less prominent low-frequency fluctuations as a function of age also in the BOLD-signal. Additionally, we found slightly higher offset values in the right hemisphere compared to the left hemisphere, indicating greater power of the BOLD signal in the right hemisphere. However, it should be noted that the difference was negligible, only 0.01. Previous studies align with our findings, suggesting earlier maturation of the right hemisphere, as cortical folding occurs earlier in the right hemisphere in fetuses. Furthermore, a general functional dominance (except for linguistic stimuli) of the right hemisphere has been described in fetuses, neonates and preterm-born infants.^64^

### Postnatal age and aperiodic parameters

Our finding of decreasing exponent values (flattening power spectrum) as a function of postnatal age is consistent with several previous EEG studies, such as a longitudinal study in infants^27^, and several cross-sectional studies during early childhood to adolescence^28,29^ and into early-adulthood.^30^ However, these associations between age and aperiodic parameters are not necessarily linear throughout the lifespan, as McSweeney et al.^34^ demonstrated quadratic age-related changes of offset and exponent values during the childhood (in 4-11 year-olds).

Our findings suggest that after birth, in the very early stages of brain maturation, the distribution of aperiodic activity (power of BOLD signal) on different frequencies changes (flattens). This could reflect the emergence of more widespread or global neural processes during aging, or as discussed above, changes in brain volume or myelination. In EEG, changes in the steepness of the slope of power spectrum have been thought to reflect cortical synaptic excitation–inhibition (E–I) balance^65^, undergoing changes from the very early stages of development onwards.^66^ In animal studies, a steeper spectral slope (higher exponent) has been correlated to reduced E-I balance in macaque and rat cortices, possibly due to greater gamma-aminobutyric acid (GABA) synapse density.^65^ The aperiodic exponent has been implicated in pathology in Attention Deficit Hyperactivity Disorder (ADHD)^67,68^ and autistic traits.^69^ An optimal E-I balance may play a crucial role in typical brain development, as medical treatment has been shown to lead to the “normalization” of E-I balance (exponent) in ADHD children.^68^ More studies examining the age-related changes of the periodic and aperiodic parameters of the fMRI BOLD signal, especially longitudinal ones, are required.

### Preterm birth and aperiodic parameters

In our study, we observed smaller exponents in preterm neonates compared to term-born neonates indicating a more flattened BOLD power distribution across frequencies. Parallel trend was seen with offsets, but the association did not reach statistical significance. Given the increasing trend in exponent values during postmenstrual aging, our findings likely reflect the shorter duration of pregnancy in preterm compared to term-born neonates. Our finding is consistent with previous fMRI BOLD signal study demonstrating altered low-frequency fluctuation amplitudes in preterm newborns, particularly in the motor and primary sensor cortices.^70^ As discussed earlier, a flattened power spectrum, at least according to prior EEG literature, may indicate a shift towards excitation, reflecting the changes in the E-I balance. Prior research has shown that preterm-born children often face delay in inhibition abilities, though most behavioral studies suggest that they catch up with their peers by late childhood.^71^ Alternatively, smaller exponents may reflect more rapid postnatal neural maturational processes in preterm versus term-born neonates. Follow-up studies are needed to investigate whether the trajectory of aperiodic parameters continues to evolve throughout the lifespan.

### Sex and aperiodic parameters

We observed smaller exponents and higher offsets in females compared to males. Sex-dependent differences in brain development have been described in previous studies, including increased functional connectivity of the visual association network in female infants^25^ and higher fractional anisotropy in females within the posterior and temporal white matter in 5-year-olds.^72^ Our findings align with these studies, tentatively suggesting faster brain maturation (stronger flattening of the power spectrum) in females. Further studies are needed to examine whether these sex-specific differences persist into later childhood.

### Machine learning based age prediction

In our machine learning based analysis, we found that both aperiodic parameters of the BOLD signal can be used to predict neonate age with moderate to relatively high performance (*R²’s* 0.20–0.41). The offset parameter generally yielded slightly higher prediction performance estimates than the exponent parameters, and using both kinds of parameters further improved the prediction performance estimates. We used machine learning to provide support for aperiodic parameters as a viable measure of brain maturation and this approach has connections to an emerging field aims to predict age from the brain images. Previously, Kardan et al.^73^, predicted infants’ postnatal age from rs-fMRI data as a regression task with high performance (*median R*^2^ = 0.51), but it should be noted that the dataset differed from that of the present study, especially in that the subjects’ ages were considerably higher and spanned a wider range (8–26 months). Majority of preceding studies in predicting neonate or infant age have been performed using structural^24,74,75^ or multi-modal^76–78^ MRI data. The few studies focusing on predicting neonate or infant age using functional MRI are mostly implemented as classification tasks^79,80^, with Smyser et al.^80^ also modeling age from gestation at term age as a regression task. Regarding regional associations between rs-fMRI and age, Dosenbach et al.^81^ found anterior prefrontal cortex and precuneus to contribute most to age predictions on older subjects between 7 to 30 years. Brain age prediction has many potential uses as means to gain insights to normal development and the brain age estimates in turn can be used as predictor for various brain conditions and disorders, to be further used in clinical context.^82,83^ Systematic bias in age estimations has been reported^84^, inviting for more comprehensive models to be used to avoid overestimation of age in young subjects.

### Limitations

It should be noted that our study design is cross-sectional and therefore no assumptions about individual changes in the pace of brain maturation can be made. However, owing to our large sample size, we are confident that our group-level findings are generalizable to a larger population. While the cross validated test performance estimates demonstrate relatively high generalization ability of the model, it should be noted that the training performances are considerably better, implying overfitting despite model regularization.^85^ Among new-born infants, sleep-wake patterns start to evolve soon after birth, first changing rapidly during the first few months after birth from relatively quiet and active sleep phases to structured non-rapid eye-movement and rapid eye-movement sleep.^86–88^ Their sleep is dispersed unevenly throughout the day^89^ and uncertainty as to the state of wakefulness or the stage of sleep at the time of scan can complicate the interpretation of fMRI.^53^ Because such effect is very difficult to avoid in this age group, we assume that changes in aperiodic parameters are related to aging, premature birth, and sex of the neonates. It has been demonstrated that functional connections strongly related to age as a target variable are more intercorrelated than expected by chance, providing redundant information ultimately resulting in unreliable weights.^90^ While our study used BOLD signal parameters as features, rather than functional connections, similar limitation may apply to the interpretability of our analysis as well. Finally, in our data set, the variation of the postnatal age in neonates was low, due to the choice of performing the scans close to birth. This led to highly skewed distribution of the infant age data. However, the visually inspected diagnostics of the linear regression models were acceptable.

### Conclusions

In this study we showed differences in aperiodic parameters of rs-fMRI BOLD signal in preterm vs term-born neonates and between males and females. We also demonstrated the applicability of those aperiodic parameters for the purpose of machine learning based neonate age prediction with relatively high performance (*R²* = 0.41) and identified the most important ROIs to that end. Based on our results, we consider the rs-fMRI BOLD signal and its aperiodic component a viable functional brain metric for quantifying brain maturation in neonates. In addition to that, we hope that our results will enable more precise surveying of the neural origins of developmental conditions and disorders.

## Acknowledgements

Data were provided by the developing Human Connectome Project, KCL-Imperial-Oxford Consortium funded by the European Research Council under the European Union Seventh Framework Programme (FP/2007-2013) / ERC Grant Agreement no. [319456]. We are grateful to the families who generously supported this trial.

## Funding

Ilkka Suuronen

- Emil Aaltonen Foundation

Silja Luotonen

- The Finnish Cultural Foundation / Varsinais-Suomi Regional Fund

Elmo P. Pulli

- Emil Aaltonen Foundation
- Juho Vainio Foundation
- Päivikki and Sakari Sohlberg Foundation
- Finnish Brain Foundation
- The Finnish Cultural Foundation
- Strategic Research Council (SRC) established within the Research Council of Finland (#352648 and subproject #352655)

Niloofar Hashempour

- University of Turku Graduate School

Jetro J. Tuulari,

- Finnish Medical Foundation
- Emil Aaltonen Foundation
- Sigrid Juselius Foundation
- Signe and Ane Gyllenberg Foundation
- Hospital District of Southwest Finland State Research Grants

Harri Merisaari

- Academy of Finland (#26080983)

## Supplementary material

Online supplemental material is available for this article.

