## Supplemental file 1 for "Aperiodic parameters of the fMRI power spectrum associate with preterm birth and neonatal age"

### Supplemental file 1: Methods

#### Linear Mixed Effect Regression Models

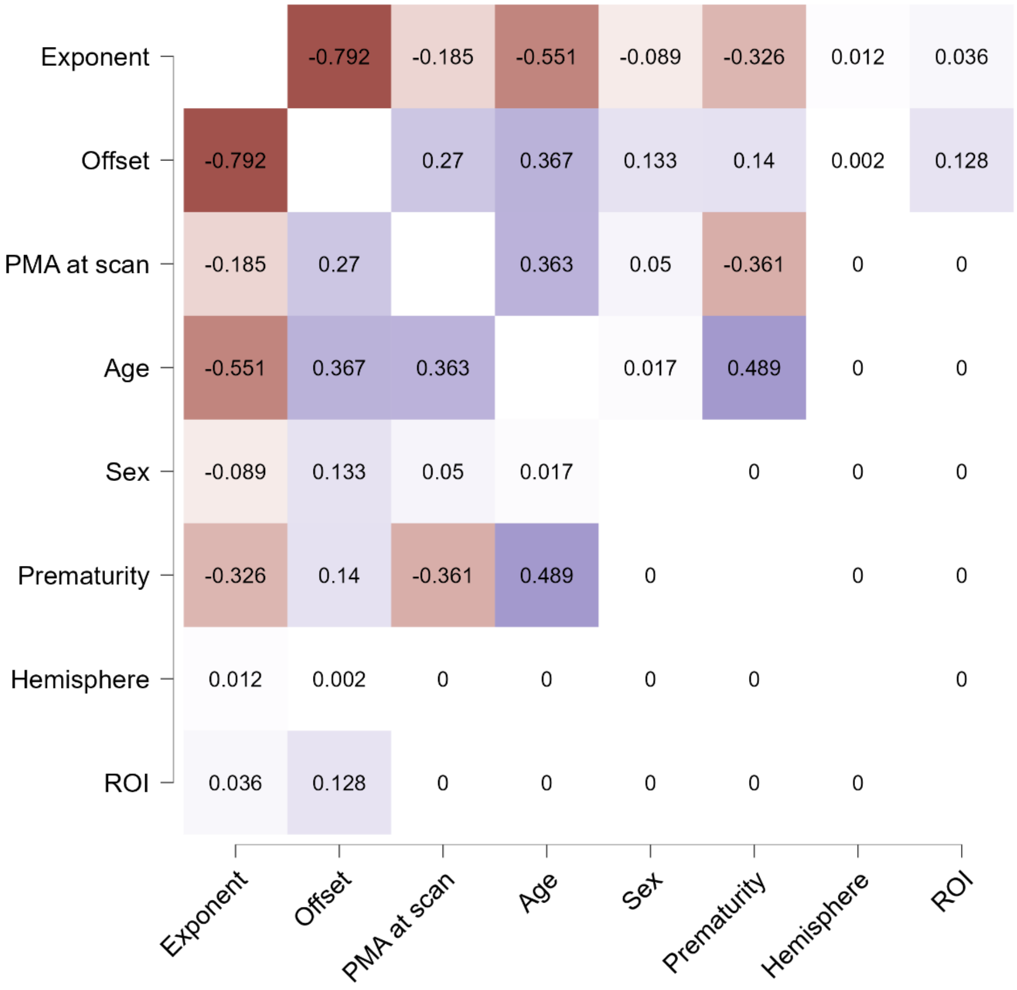


**Figure S1.** Correlation matrix (Spearman correlation) for the main variables of interest, conditioned on outliers based on DVARS. *PMA at scan* = postmenstrual age (PMA) of the neonate at scan (weeks), *Age* = postnatal age (age from birth; weeks), *Sex* = biological sex of the child (male = 0, female = 1), *Prematurity* = term versus preterm birth (term-born = 0, preterm = 1; postmenstrual age at birth < 37 weeks), *Hemisphere* = left (= 0) or right (= 1) side of the brain and *ROI* = postcentral (= 0) or precentral (= 1) gyri.

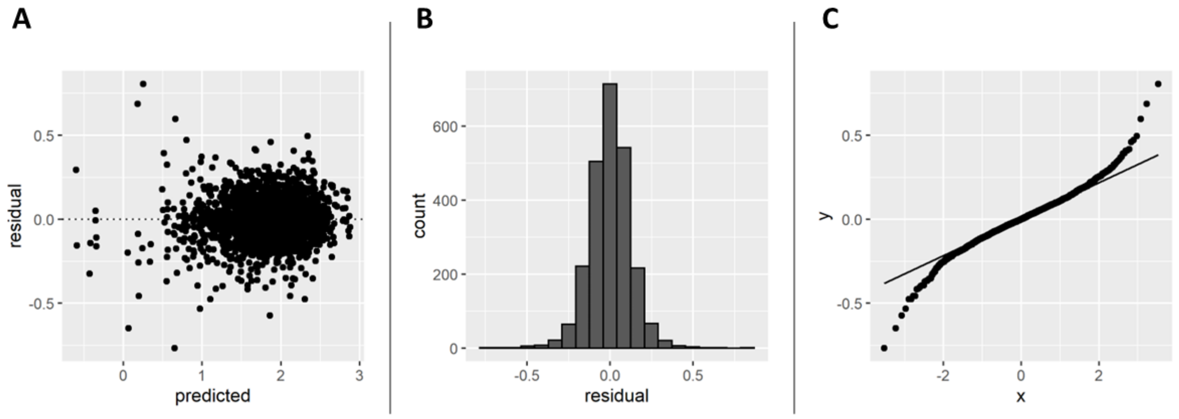


**Figure S2.** Model diagnostics for Model 1 (Exponent) before excluding residual outliers. A) Predicted values vs. residuals. B) Histogram of the residual distribution. C) Q-Q plot of the residuals.

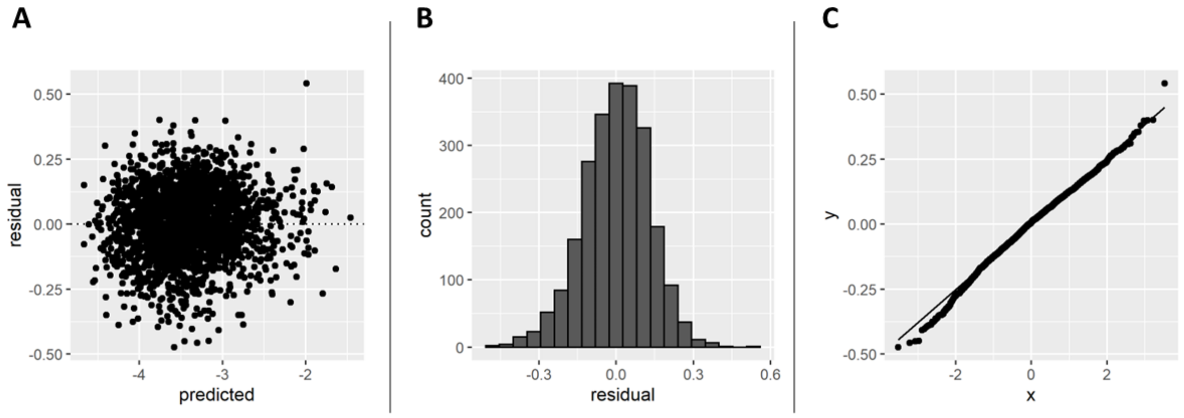


**Figure S3.** Model diagnostics for Model 2 (Offset). A) Predicted values vs. residuals. B) Histogram of the residual distribution. C) Q-Q plot of the residuals.

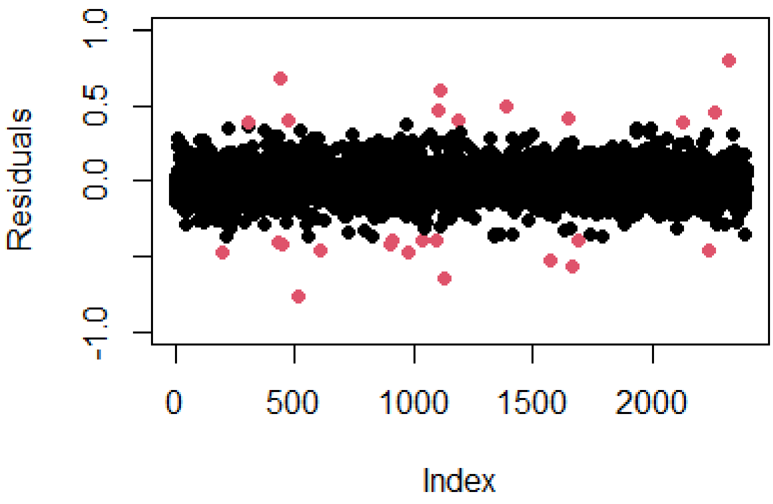


**Figure S4.** Outliers (± 3 SD) of the residuals from Model 1 (Exponent). Outliers (N = 26) marked with red color.

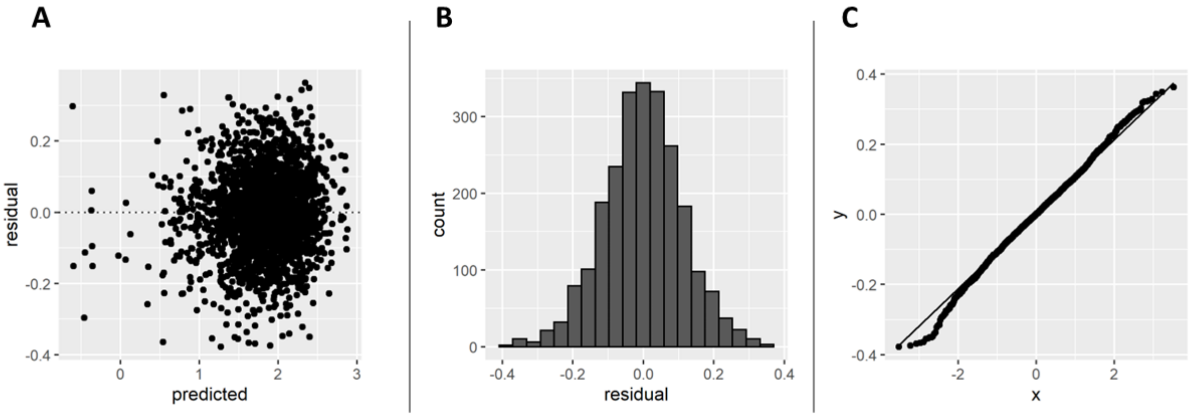


**Figure S5.** Model diagnostics for Model 1 (Exponent) after excluding residual outliers (N = 26). A) Predicted values vs. residuals. B) Histogram of the residual distribution. C) Q-Q plot of the residuals.

**Table S1.** Results of the linear mixed-effects regression model for exponent (Model 1). Residual outliers (N = 26) not excluded.

| | Estimate | SE | df | t value | p value |
|---|---|---|---|---|---|
| Intercept | 1.89 | 0.03 | 651 | 69.43 | 0.000*** |
| PMA at scan | 0.02 | 0.02 | 611 | 0.94 | 0.348 |
| ROI | 0.02 | 0.01 | 1797 | 1.89 | 0.060 |
| Postnatal age | -0.17 | 0.03 | 619 | -5.95 | 0.000*** |
| Sex | -0.09 | 0.03 | 641 | -2.70 | 0.007** |
| Prematurity | -0.18 | 0.08 | 639 | -2.19 | 0.029* |
| Hemisphere | 0.01 | 0.01 | 1797 | 1.59 | 0.112 |
| PMA at scan : ROI | -0.03 | 0.01 | 1797 | -3.55 | 0.000*** |
| Postnatal age : ROI | -0.02 | 0.01 | 1797 | -1.54 | 0.123 |
| Sex : ROI | 0.01 | 0.01 | 1797 | 0.92 | 0.358 |
| Prematurity : ROI | 0.03 | 0.03 | 1797 | 1.15 | 0.249 |

**Table S2.** Comparing results of Model 1 (exponent) before vs. after excluding residual outliers. Reporting differences between coefficients (modified model – original model) and sensitivity (((modified model – original model) / original model) *100) of the coefficients.

| | Estimate | SE | df | t value | p value |
|---|---|---|---|---|---|
| Intercept | 0.002 (0.10 %) | 0,000 (- 0.14 %) | -11.352 (- 1.74) | 0.164 (0.24 %) | 0.000 (32 666.87 %) |
| PMA at scan | -0.002 (- 7.08 %) | 0,000 (0.30 %) | -0.006 (0.00 %) | -0.069 (- 7.37 %) | 0.037 (10.51 %) |
| ROI | -0.004 (- 20.63 %) | -0.001 (-9.35 %) | -26.454 (- 1.47 %) | -0.235 (-12.44 %) | 0.039 (66.19 %) |
| Postnatal age | -0.003 (1.47 %) | 0.000 (0.03 %) | -7.726 (- 1.25 %) | -0.085 (1.43 %) | 0.000 (-38.73) |
| Sex | -0.002 (2.05 %) | 0.000 (- 0.03 %) | -8.807 (- 1.37 %) | -0.056 (2.08%) | -0.001 (-15.51%) |
| Prematurity | 0.007 (- 3.77 %) | 0.000 (0.01 %) | -8.095 (- 1.27 %) | 0.083 (- 3.78) | 0.007 (22.84 %) |
| Hemisphere | -0.001 (-5.65 %) | -0.001 (- 9.58 %) | -26.818 (- 1.49 %) | 0.069 (4.34 %) | -0.015 (-13.13 %) |
| PMA at scan : ROI | 0.005 (-17.52 %) | -0.001 (- 6.06 %) | -21.474 (- 1.19 %) | 0.433 (-12.20 %) | 0.001 (368.97 %) |
| Postnatal age : ROI | 0.004 | -0.001 | -25.136 | 0.282 | 0.085 |

| | | | | | |
|---|---|---|---|---|---|
| | (-24.69 %) | (- 7.82 %) | (- 1.40 %) | (-18.30 %) | (68.76 %) |
| Sex : ROI | 0.005 (41.47 %) | -0.001 (- 9.56 %) | -26.742 (- 1.49 %) | 0.519 (56.41 %) | -0.207 (-57.98 %) |
| Prematurity : ROI | -0.006 (-17.40 %) | -0.003 (- 9.04 %) | -25.878 (- 1.44 %) | -0.106 (-9.19 %) | 0.046 (18.53 %) |

#### Linear Regression Models

To map the estimates of *Prematurity* (term-born = 0, preterm = 1) for aperiodic parameters (exponent or offset) of fMRI BOLD signal from 90 different brain areas, the *lm* function from *stats* package were used. The formula of models followed the formulas of the linear mixed effect regression models described above:

Model 1: *Exponent ~ PMA at scan + Age + Sex + Prematurity + Motion*

Model 2: *Offset ~ PMA at scan + Age + Sex + Prematurity + Motion + Exponent*

where *PMA at scan* means the postmenstrual age (PMA) of the neonate at scan (weeks; z-transformed) and *Age* means the postnatal age of the child (weeks; z-transformed). *Sex* means the sex of the child (male = 0, female = 1) and *Prematurity* means term versus preterm birth (term-born = 0, preterm = 1; postmenstrual age at birth < 37 weeks). *Motion* means the number of motion-compromised volumes (outliers based on DVARS; z-transformed). In Model 2, the exponent was z-transformed. P-values were corrected by FDR correction using the Benjamini-Hochberg method.
