## Supplemental file 2 for "Aperiodic parameters of the fMRI power spectrum associate with preterm birth and neonatal age"

### Supplemental file 2: Results

#### Linear Mixed Effect Regression Models

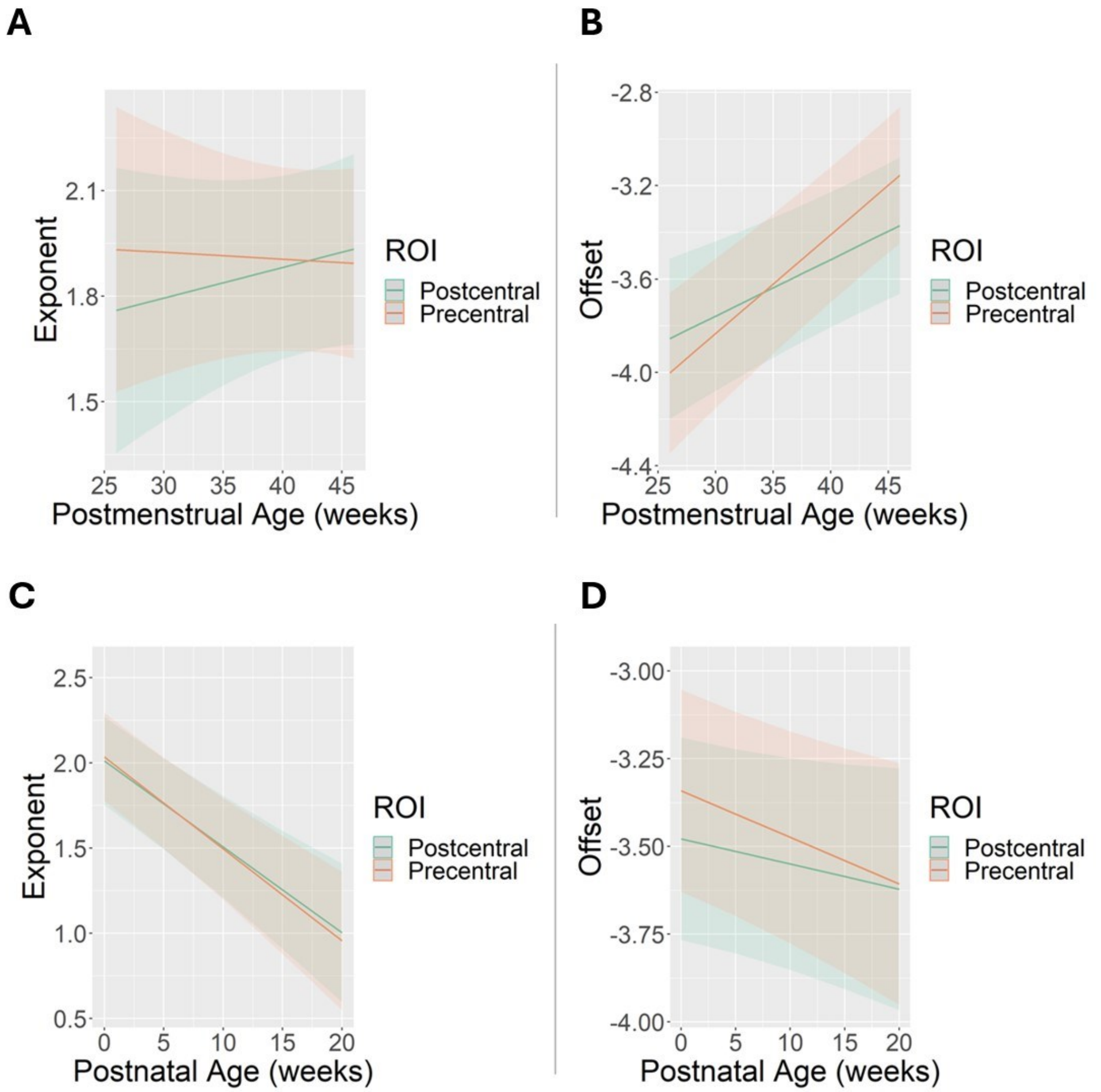


**Figure S1.** A) Postmenstrual age (weeks) and predicted aperiodic exponents (conditioned on random effects) from the pre- and postcentral gyri. B) Postmenstrual age (weeks) and predicted aperiodic offsets (conditioned on random effects) from the pre- and postcentral gyri. C) Postnatal age (weeks) and predicted aperiodic exponents from the pre- and postcentral gyri. D) Postnatal age (weeks) and predicted aperiodic offsets (conditioned on random effects) from the pre- and postcentral gyri. ROI = region-of-interest.

### Linear Regression Models

**Table S1.** Estimates, p-values and FDR corrected p-values of *Prematurity* from linear regression model (exponent). Statistically significant results after FDR correction marked with grey color.

| Brain area | Estimate | p value | Adjusted p value |
|---|---|---|---|
| Precentral_L | -0.14 | 0.103 | 0.171 |
| Precentral_R | -0.12 | 0.155 | 0.218 |
| Frontal_Sup_L | -0.09 | 0.320 | 0.379 |
| Frontal_Sup_R | -0.18 | 0.046 | 0.125 |
| Frontal_Sup_Orb_L | -0.13 | 0.138 | 0.200 |
| Frontal_Sup_Orb_R | -0.05 | 0.597 | 0.656 |
| Frontal_Mid_L | -0.03 | 0.734 | 0.769 |
| Frontal_Mid_R | -0.03 | 0.697 | 0.738 |
| Frontal_Mid_Orb_L | -0.23 | 0.006 | 0.064 |
| Frontal_Mid_Orb_R | -0.18 | 0.040 | 0.115 |
| Frontal_Inf_Oper_L | -0.18 | 0.077 | 0.150 |
| Frontal_Inf_Oper_R | -0.21 | 0.020 | 0.101 |
| Frontal_Inf_Tri_L | -0.07 | 0.477 | 0.551 |
| Frontal_Inf_Tri_R | -0.20 | 0.031 | 0.102 |
| Frontal_Inf_Orb_L | -0.21 | 0.025 | 0.101 |
| Frontal_Inf_Orb_R | -0.26 | 0.004 | 0.060 |
| Rolandic_Oper_L | -0.16 | 0.067 | 0.140 |
| Rolandic_Oper_R | -0.16 | 0.051 | 0.128 |
| Supp_Motor_Area_L | -0.17 | 0.060 | 0.137 |
| Supp_Motor_Area_R | -0.15 | 0.090 | 0.165 |
| Olfactory_L | -0.14 | 0.080 | 0.154 |
| Olfactory_R | -0.23 | 0.005 | 0.063 |
| Frontal_Sup_Medial_L | -0.18 | 0.064 | 0.138 |
| Frontal_Sup_Medial_R | -0.17 | 0.065 | 0.138 |
| Frontal_Med_Orb_L | -0.24 | 0.010 | 0.092 |
| Frontal_Med_Orb_R | -0.20 | 0.026 | 0.101 |
| Rectus_L | -0.14 | 0.107 | 0.172 |
| Rectus_R | -0.19 | 0.026 | 0.101 |
| Insula_L | -0.21 | 0.029 | 0.102 |
| Insula_R | -0.16 | 0.085 | 0.159 |
| Cingulum_Ant_L | -0.24 | 0.015 | 0.101 |
| Cingulum_Ant_R | -0.22 | 0.019 | 0.101 |
| Cingulum_Mid_L | -0.14 | 0.099 | 0.169 |
| Cingulum_Mid_R | -0.19 | 0.028 | 0.102 |
| Cingulum_Post_L | -0.11 | 0.266 | 0.346 |
| Cingulum_Post_R | -0.19 | 0.035 | 0.109 |
| Hippocampus_L | -0.23 | 0.003 | 0.060 |
| Hippocampus_R | -0.31 | 0.000 | 0.009 |
| ParaHippocampal_L | -0.21 | 0.017 | 0.101 |
| ParaHippocampal_R | -0.28 | 0.001 | 0.029 |
| Amygdala_L | -0.04 | 0.549 | 0.618 |
| Amygdala_R | -0.07 | 0.302 | 0.367 |

| | | | |
|---|---|---|---|
| Calcarine_L | -0.16 | 0.064 | 0.138 |
| Calcarine_R | -0.13 | 0.120 | 0.179 |
| Cuneus_L | -0.14 | 0.121 | 0.179 |
| Cuneus_R | -0.23 | 0.006 | 0.064 |
| Lingual_L | -0.21 | 0.022 | 0.101 |
| Lingual_R | -0.17 | 0.053 | 0.129 |
| Occipital_Sup_L | -0.13 | 0.107 | 0.172 |
| Occipital_Sup_R | -0.19 | 0.024 | 0.101 |
| Occipital_Mid_L | -0.09 | 0.296 | 0.367 |
| Occipital_Mid_R | -0.04 | 0.632 | 0.685 |
| Occipital_Inf_L | -0.06 | 0.484 | 0.552 |
| Occipital_Inf_R | -0.17 | 0.047 | 0.125 |
| Fusiform_L | -0.15 | 0.095 | 0.166 |
| Fusiform_R | -0.26 | 0.004 | 0.060 |
| Postcentral_L | -0.14 | 0.093 | 0.166 |
| Postcentral_R | -0.19 | 0.024 | 0.101 |
| Parietal_Sup_L | -0.04 | 0.644 | 0.690 |
| Parietal_Sup_R | -0.12 | 0.158 | 0.218 |
| Parietal_Inf_L | 0.00 | 0.980 | 0.991 |
| Parietal_Inf_R | -0.19 | 0.030 | 0.102 |
| SupraMarginal_L | -0.14 | 0.096 | 0.166 |
| SupraMarginal_R | -0.16 | 0.070 | 0.143 |
| Angular_L | -0.10 | 0.283 | 0.358 |
| Angular_R | -0.14 | 0.109 | 0.172 |
| Precuneus_L | -0.18 | 0.037 | 0.110 |
| Precuneus_R | -0.24 | 0.004 | 0.060 |
| Paracentral_Lobule_L | -0.13 | 0.145 | 0.208 |
| Paracentral_Lobule_R | -0.13 | 0.169 | 0.230 |
| Caudate_L | -0.14 | 0.074 | 0.148 |
| Caudate_R | -0.13 | 0.121 | 0.179 |
| Putamen_L | -0.13 | 0.112 | 0.174 |
| Putamen_R | -0.09 | 0.298 | 0.367 |
| Pallidum_L | -0.08 | 0.306 | 0.367 |
| Pallidum_R | -0.09 | 0.269 | 0.346 |
| Thalamus_L | 0.00 | 0.960 | 0.982 |
| Thalamus_R | -0.17 | 0.050 | 0.127 |
| Heschl_L | -0.13 | 0.185 | 0.244 |
| Heschl_R | -0.22 | 0.020 | 0.101 |
| Temporal_Sup_L | -0.01 | 0.913 | 0.945 |
| Temporal_Sup_R | -0.12 | 0.176 | 0.237 |
| Temporal_Pole_Sup_L | -0.22 | 0.016 | 0.101 |
| Temporal_Pole_Sup_R | -0.17 | 0.044 | 0.123 |
| Temporal_Mid_L | 0.00 | 0.992 | 0.992 |
| Temporal_Mid_R | -0.08 | 0.328 | 0.384 |
| Temporal_Pole_Mid_L | -0.20 | 0.016 | 0.101 |
| Temporal_Pole_Mid_R | -0.16 | 0.057 | 0.136 |
| Temporal_Inf_L | -0.19 | 0.034 | 0.108 |
| Temporal_Inf_R | -0.05 | 0.585 | 0.650 |

**Table S2.** Estimates, p-values and FDR corrected p-values of *Prematurity* from linear regression model offset). No statistically significant results after FDR correction.

| Brain area | Estimate | p value | Adjusted p value |
|---|---|---|---|
| Precentral_L | -0.02 | 0.725 | 0.972 |
| Precentral_R | -0.02 | 0.690 | 0.972 |
| Frontal_Sup_L | -0.04 | 0.454 | 0.972 |
| Frontal_Sup_R | -0.07 | 0.177 | 0.972 |
| Frontal_Sup_Orb_L | 0.01 | 0.840 | 0.972 |
| Frontal_Sup_Orb_R | 0.00 | 0.967 | 0.995 |
| Frontal_Mid_L | -0.09 | 0.124 | 0.972 |
| Frontal_Mid_R | -0.09 | 0.167 | 0.972 |
| Frontal_Mid_Orb_L | -0.06 | 0.359 | 0.972 |
| Frontal_Mid_Orb_R | -0.06 | 0.409 | 0.972 |
| Frontal_Inf_Oper_L | 0.02 | 0.650 | 0.972 |
| Frontal_Inf_Oper_R | -0.04 | 0.432 | 0.972 |
| Frontal_Inf_Tri_L | -0.04 | 0.494 | 0.972 |
| Frontal_Inf_Tri_R | -0.02 | 0.697 | 0.972 |
| Frontal_Inf_Orb_L | -0.03 | 0.503 | 0.972 |
| Frontal_Inf_Orb_R | 0.01 | 0.821 | 0.972 |
| Rolandic_Oper_L | 0.01 | 0.825 | 0.972 |
| Rolandic_Oper_R | 0.02 | 0.684 | 0.972 |
| Supp_Motor_Area_L | -0.03 | 0.526 | 0.972 |
| Supp_Motor_Area_R | 0.00 | 0.995 | 0.995 |
| Olfactory_L | -0.04 | 0.302 | 0.972 |
| Olfactory_R | -0.02 | 0.695 | 0.972 |
| Frontal_Sup_Medial_L | -0.08 | 0.120 | 0.972 |
| Frontal_Sup_Medial_R | -0.04 | 0.450 | 0.972 |
| Frontal_Med_Orb_L | 0.03 | 0.588 | 0.972 |
| Frontal_Med_Orb_R | 0.05 | 0.350 | 0.972 |
| Rectus_L | 0.03 | 0.492 | 0.972 |
| Rectus_R | 0.04 | 0.315 | 0.972 |
| Insula_L | -0.03 | 0.567 | 0.972 |
| Insula_R | -0.01 | 0.851 | 0.972 |
| Cingulum_Ant_L | -0.06 | 0.172 | 0.972 |
| Cingulum_Ant_R | -0.02 | 0.710 | 0.972 |
| Cingulum_Mid_L | -0.02 | 0.684 | 0.972 |
| Cingulum_Mid_R | 0.03 | 0.534 | 0.972 |
| Cingulum_Post_L | -0.01 | 0.870 | 0.972 |
| Cingulum_Post_R | 0.03 | 0.448 | 0.972 |
| Hippocampus_L | -0.01 | 0.842 | 0.972 |
| Hippocampus_R | 0.03 | 0.280 | 0.972 |
| ParaHippocampal_L | -0.02 | 0.500 | 0.972 |
| ParaHippocampal_R | 0.03 | 0.276 | 0.972 |
| Amygdala_L | 0.01 | 0.854 | 0.972 |
| Amygdala_R | 0.02 | 0.588 | 0.972 |
| Calcarine_L | -0.07 | 0.239 | 0.972 |
| Calcarine_R | -0.04 | 0.482 | 0.972 |
| Cuneus_L | 0.01 | 0.836 | 0.972 |

| | | | |
|---|---|---|---|
| Cuneus_R | 0.01 | 0.875 | 0.972 |
| Lingual_L | -0.02 | 0.630 | 0.972 |
| Lingual_R | -0.07 | 0.177 | 0.972 |
| Occipital_Sup_L | -0.07 | 0.203 | 0.972 |
| Occipital_Sup_R | 0.07 | 0.244 | 0.972 |
| Occipital_Mid_L | -0.12 | 0.049 | 0.972 |
| Occipital_Mid_R | 0.00 | 0.948 | 0.993 |
| Occipital_Inf_L | -0.03 | 0.548 | 0.972 |
| Occipital_Inf_R | -0.13 | 0.073 | 0.972 |
| Fusiform_L | 0.00 | 0.909 | 0.977 |
| Fusiform_R | 0.00 | 0.912 | 0.977 |
| Postcentral_L | -0.07 | 0.153 | 0.972 |
| Postcentral_R | -0.10 | 0.058 | 0.972 |
| Parietal_Sup_L | -0.11 | 0.055 | 0.972 |
| Parietal_Sup_R | -0.04 | 0.519 | 0.972 |
| Parietal_Inf_L | -0.06 | 0.274 | 0.972 |
| Parietal_Inf_R | -0.01 | 0.866 | 0.972 |
| SupraMarginal_L | 0.01 | 0.828 | 0.972 |
| SupraMarginal_R | -0.03 | 0.612 | 0.972 |
| Angular_L | -0.08 | 0.180 | 0.972 |
| Angular_R | 0.07 | 0.263 | 0.972 |
| Precuneus_L | -0.03 | 0.593 | 0.972 |
| Precuneus_R | -0.01 | 0.886 | 0.972 |
| Paracentral_Lobule_L | -0.02 | 0.755 | 0.972 |
| Paracentral_Lobule_R | -0.01 | 0.847 | 0.972 |
| Caudate_L | -0.02 | 0.484 | 0.972 |
| Caudate_R | 0.00 | 0.990 | 0.995 |
| Putamen_L | 0.01 | 0.762 | 0.972 |
| Putamen_R | 0.04 | 0.282 | 0.972 |
| Pallidum_L | -0.02 | 0.629 | 0.972 |
| Pallidum_R | 0.03 | 0.356 | 0.972 |
| Thalamus_L | -0.01 | 0.796 | 0.972 |
| Thalamus_R | 0.05 | 0.182 | 0.972 |
| Heschl_L | 0.00 | 0.926 | 0.981 |
| Heschl_R | 0.05 | 0.258 | 0.972 |
| Temporal_Sup_L | -0.08 | 0.168 | 0.972 |
| Temporal_Sup_R | -0.01 | 0.869 | 0.972 |
| Temporal_Pole_Sup_L | 0.00 | 0.974 | 0.995 |
| Temporal_Pole_Sup_R | -0.01 | 0.834 | 0.972 |
| Temporal_Mid_L | -0.08 | 0.172 | 0.972 |
| Temporal_Mid_R | 0.02 | 0.702 | 0.972 |
| Temporal_Pole_Mid_L | -0.03 | 0.509 | 0.972 |
| Temporal_Pole_Mid_R | 0.01 | 0.786 | 0.972 |
| Temporal_Inf_L | 0.01 | 0.834 | 0.972 |
| Temporal_Inf_R | 0.01 | 0.829 | 0.972 |

### Machine learning results

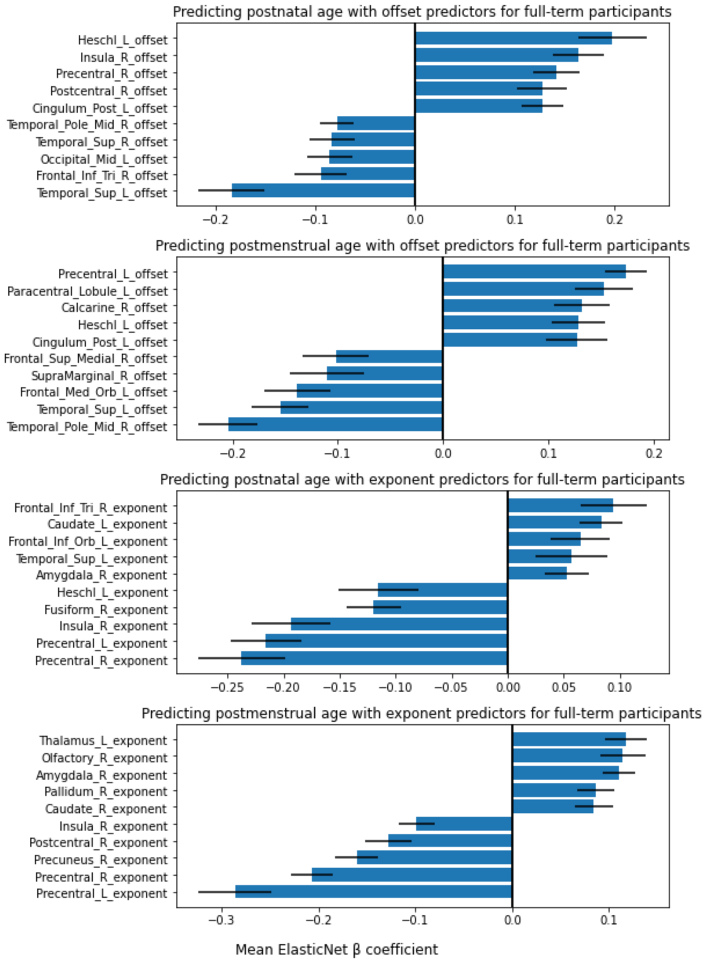


**Figure S2.** Highest positive and negative model beta coefficients for predicting neonate age in different settings on term born participants.

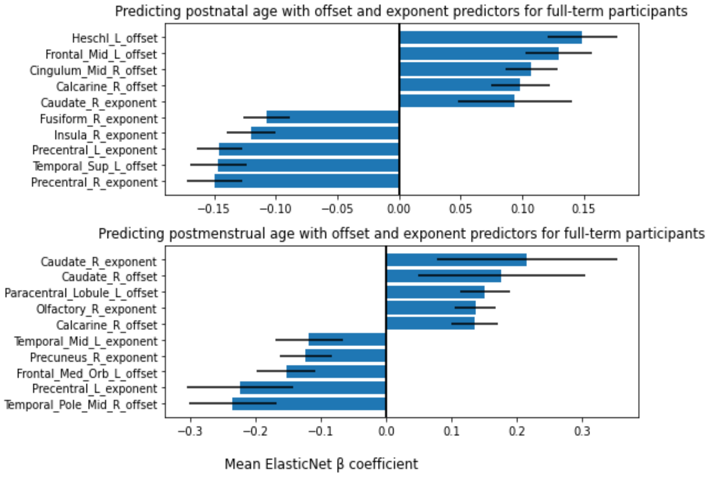


**Figure S3**. Highest positive and negative model beta coefficients for predicting neonate postnatal and postmenstrual age with both offset and exponent predictors for term born participants.

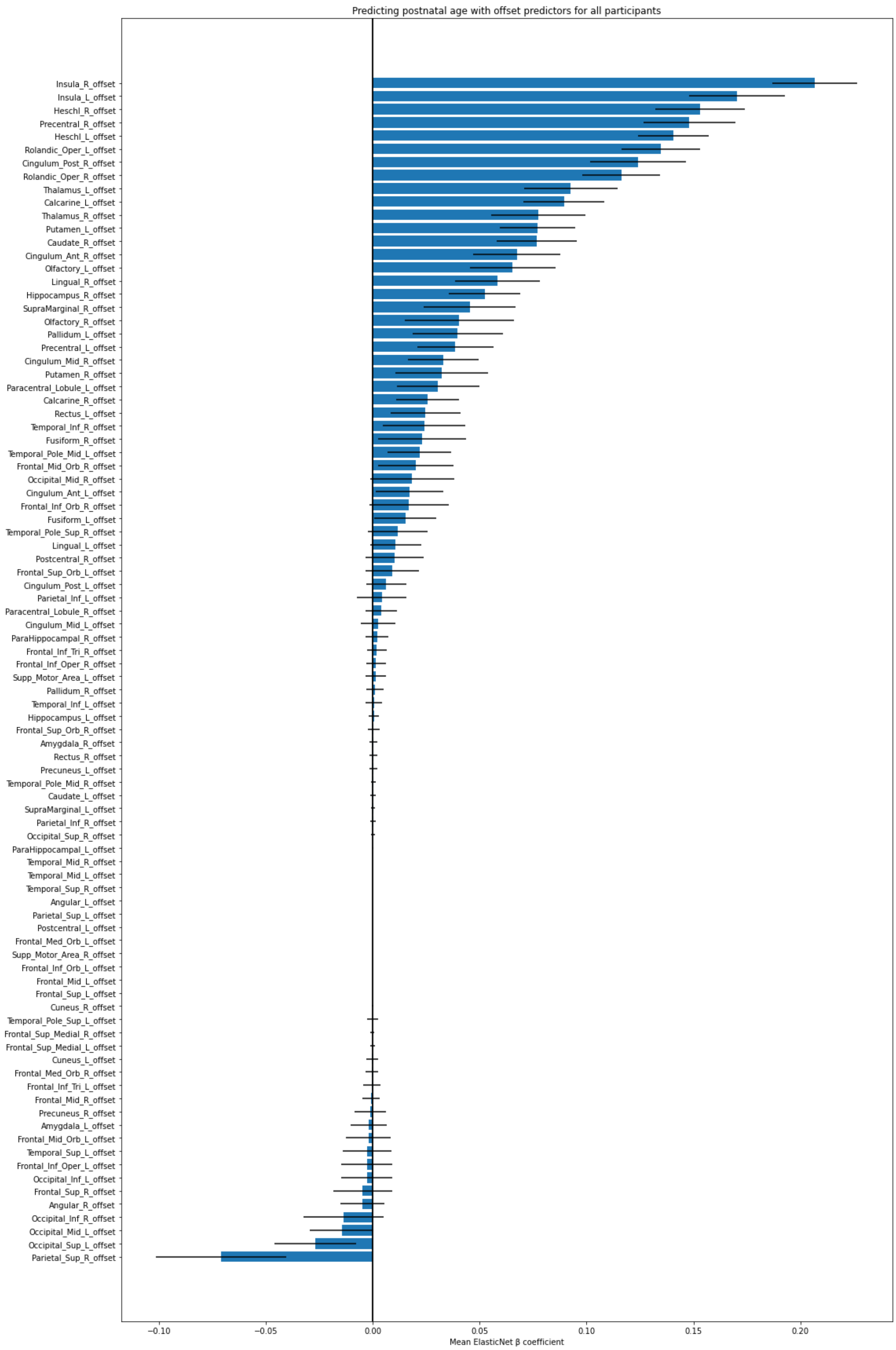


**Figure S4.** Offset feature mean coefficients for predicting postnatal age at scan. All subjects.

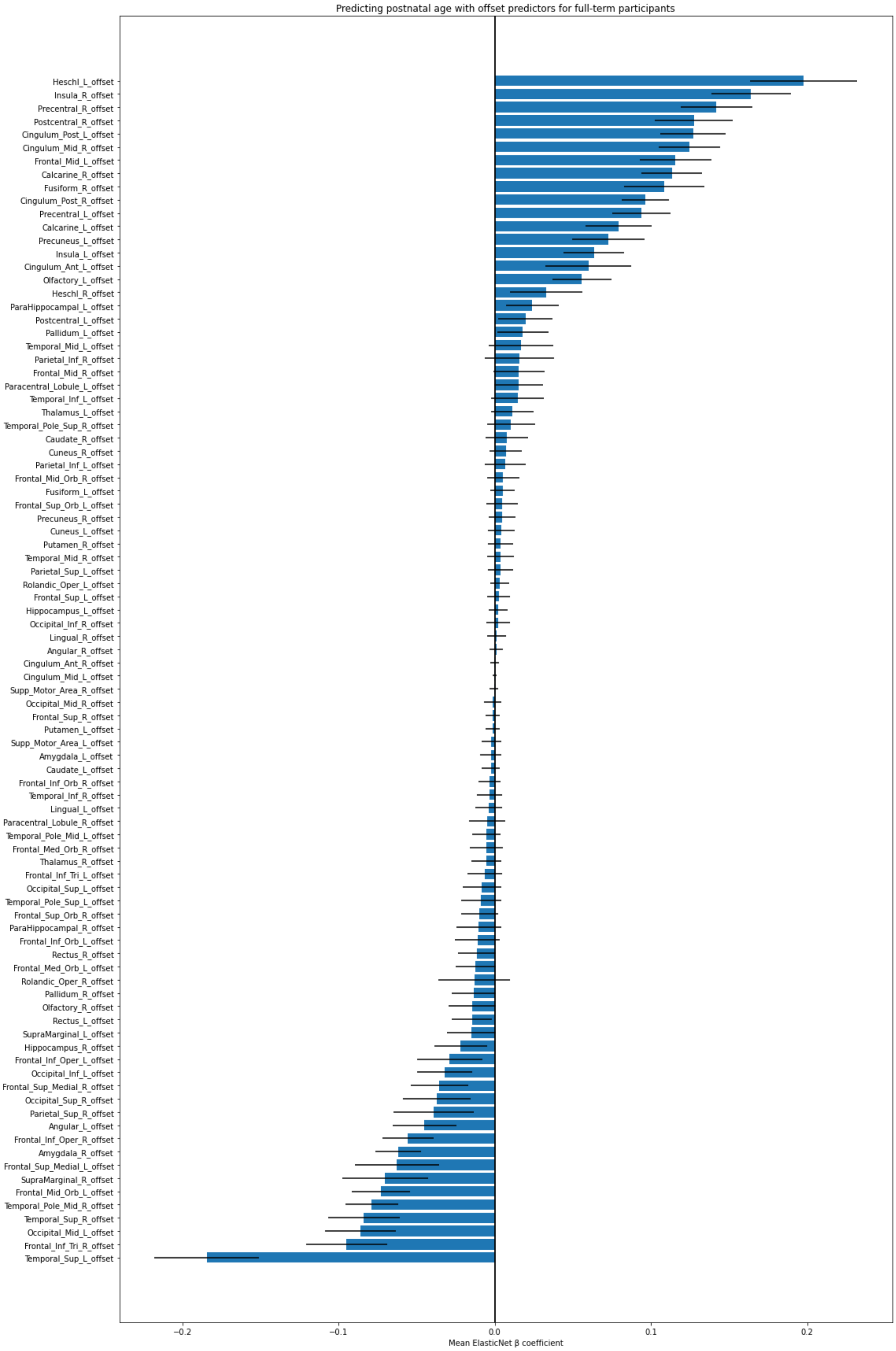


**Figure S5.** Offset feature mean coefficients for predicting postnatal age at scan. Full-term subjects.

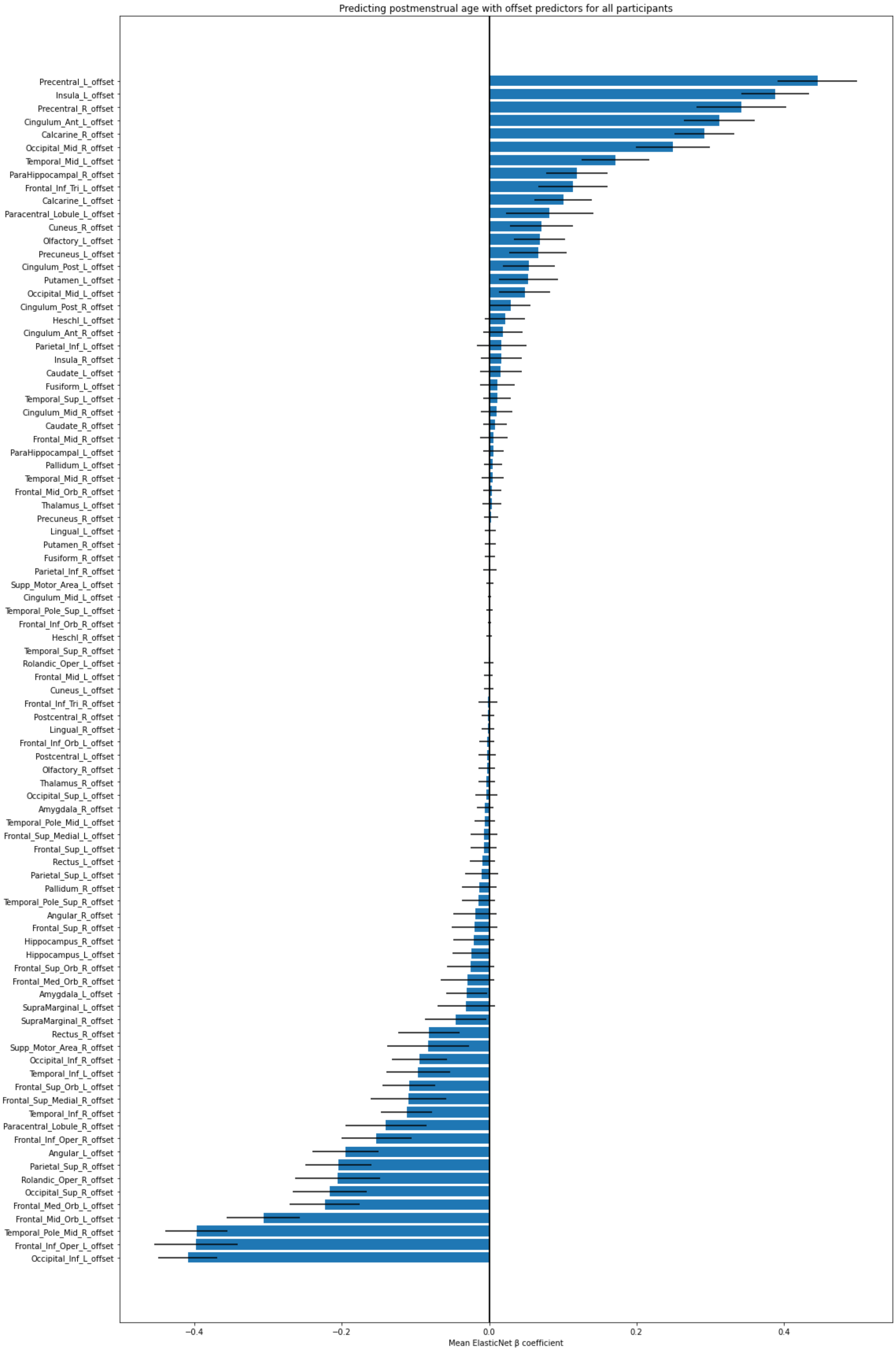


**Figure S6.** Offset feature mean coefficients for predicting postmenstrual age at scan. All subjects.

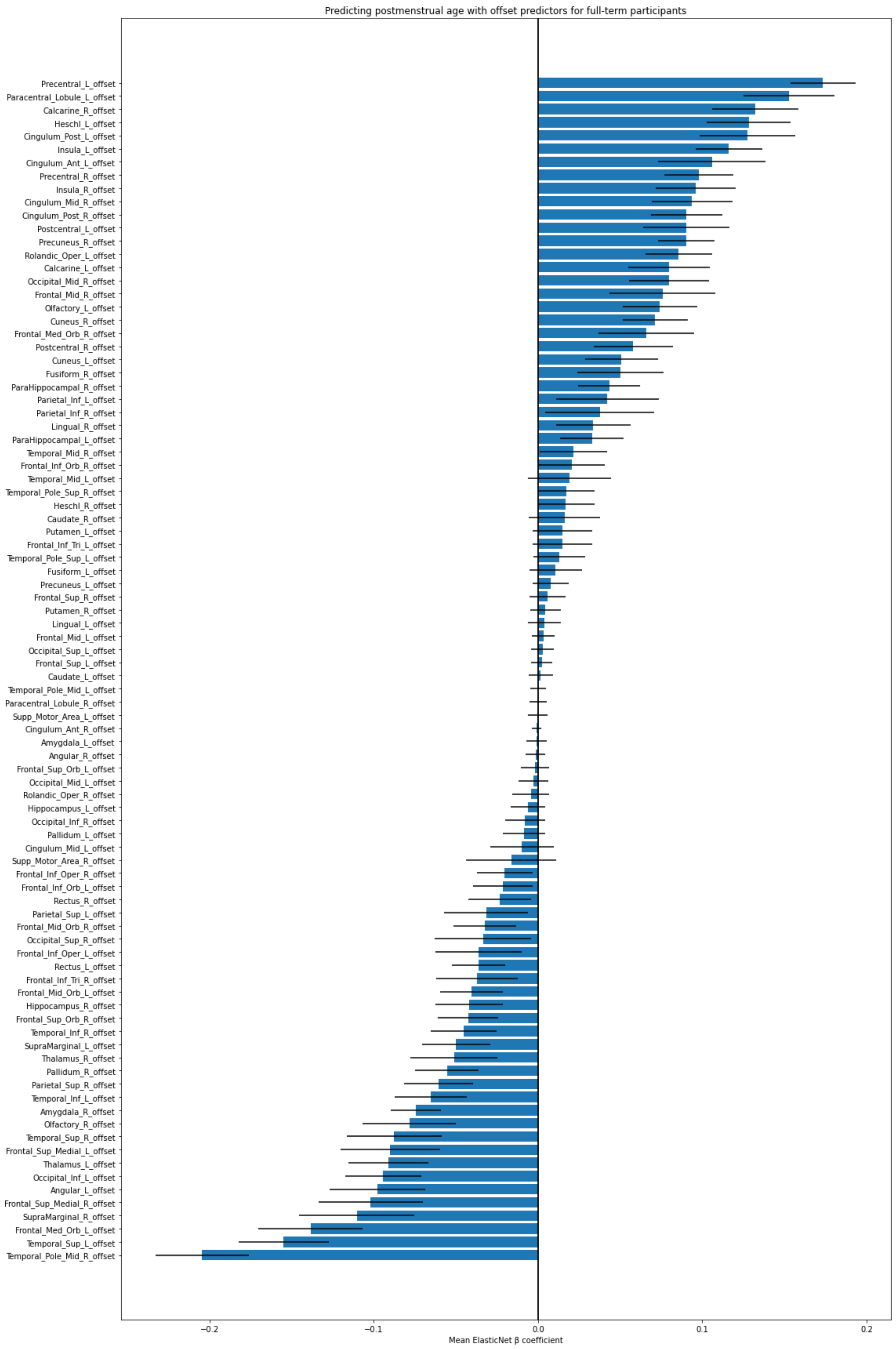


**Figure S7.** Offset feature mean coefficients for predicting postmenstrual age at scan. Full-term subjects.

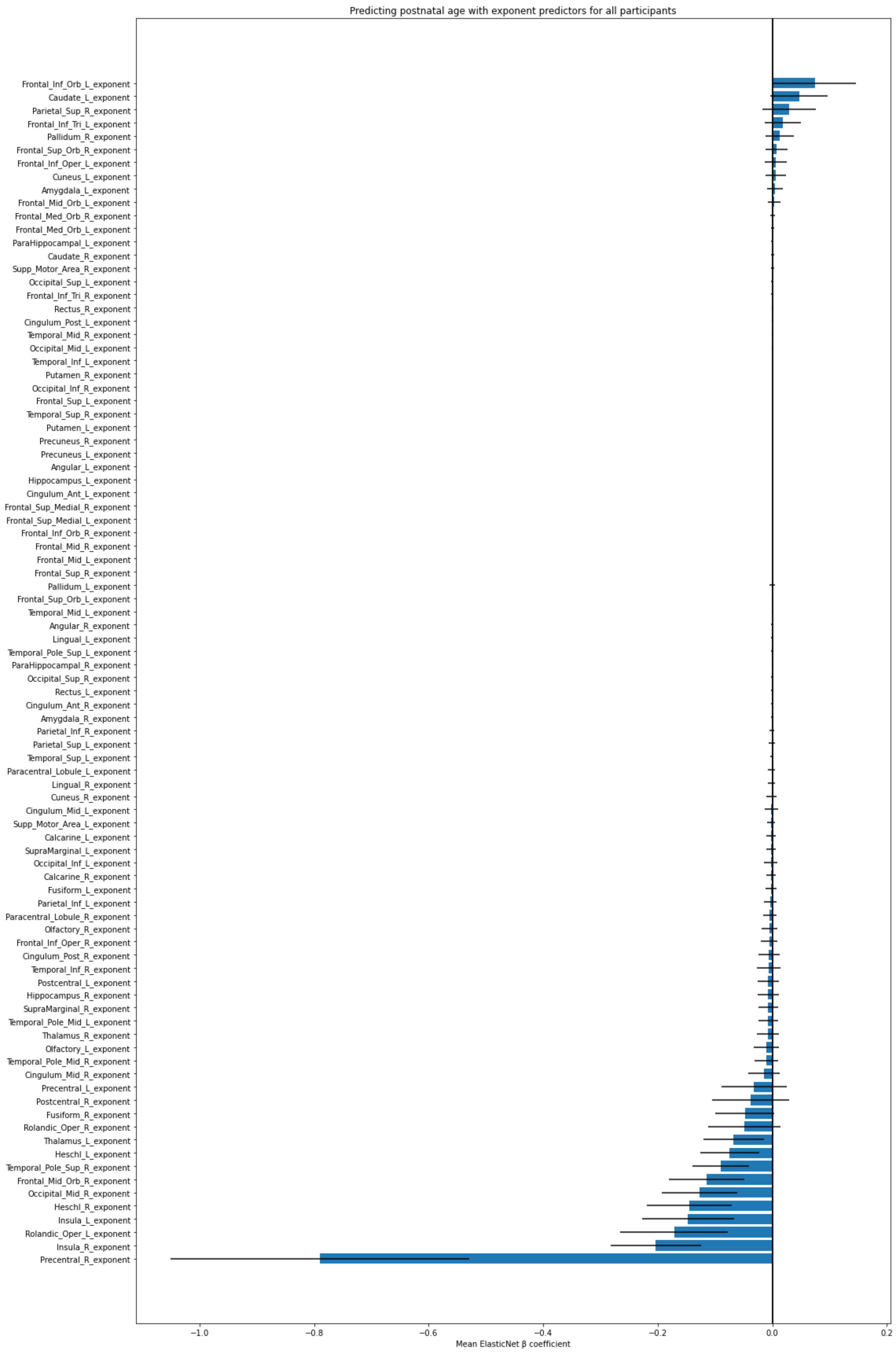


**Figure S8.** Exponent feature mean coefficients for predicting postnatal age at scan. All subjects.

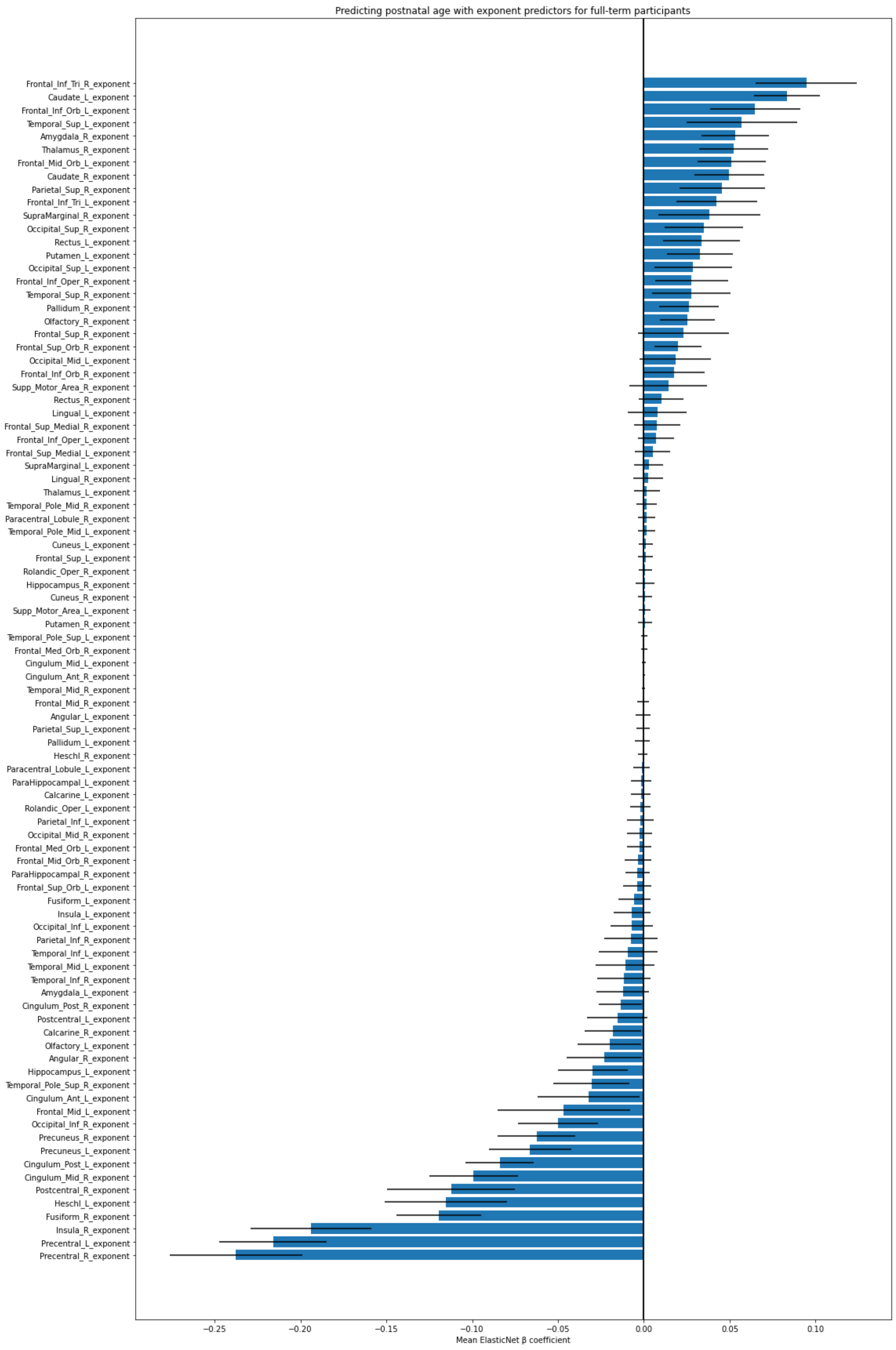


**Figure S9.** Exponent feature mean coefficients for predicting postnatal age at scan. Full-term subjects.

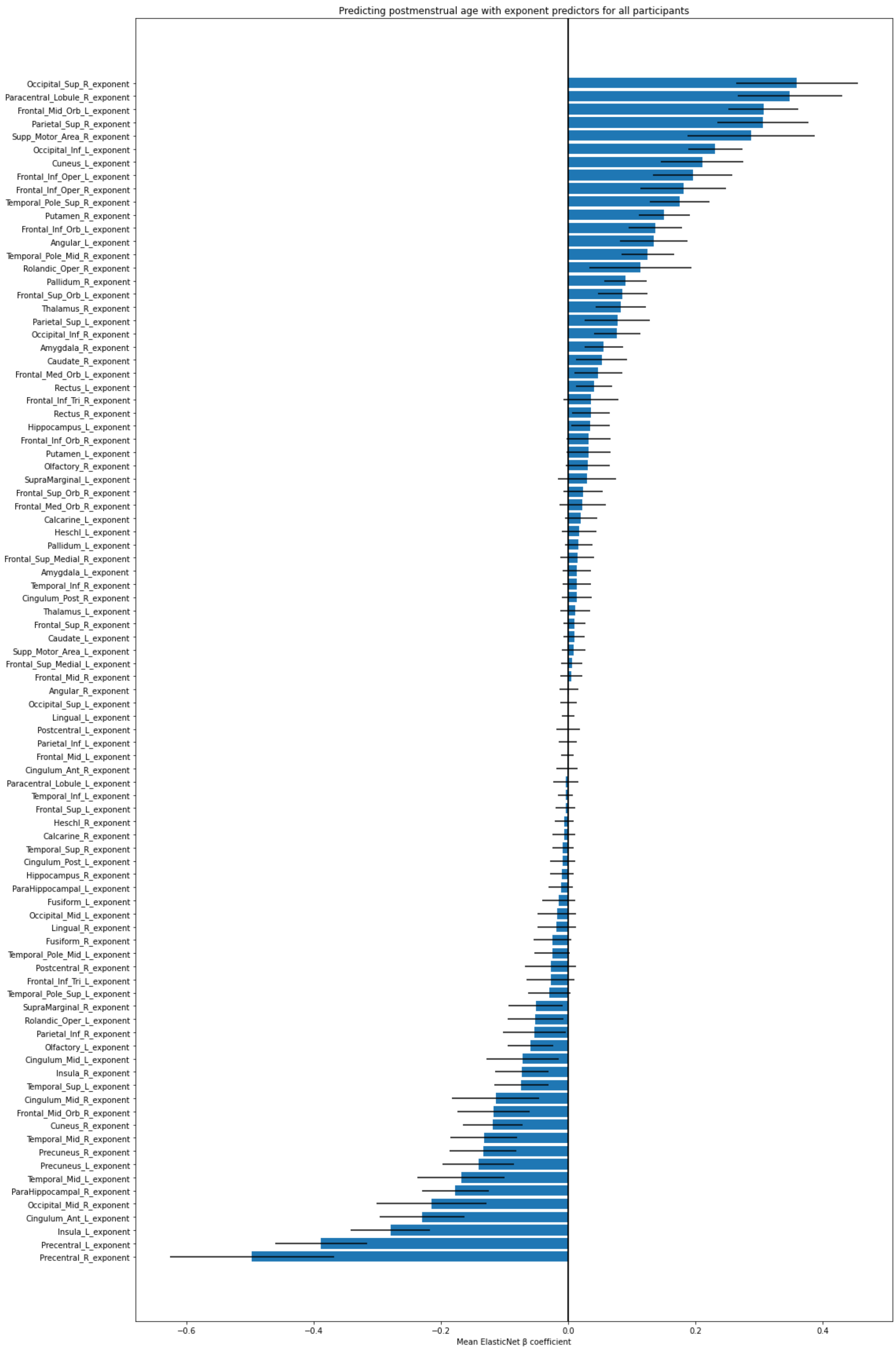


**Figure S10.** Exponent feature mean coefficients for predicting postmenstrual age at scan. All subjects.

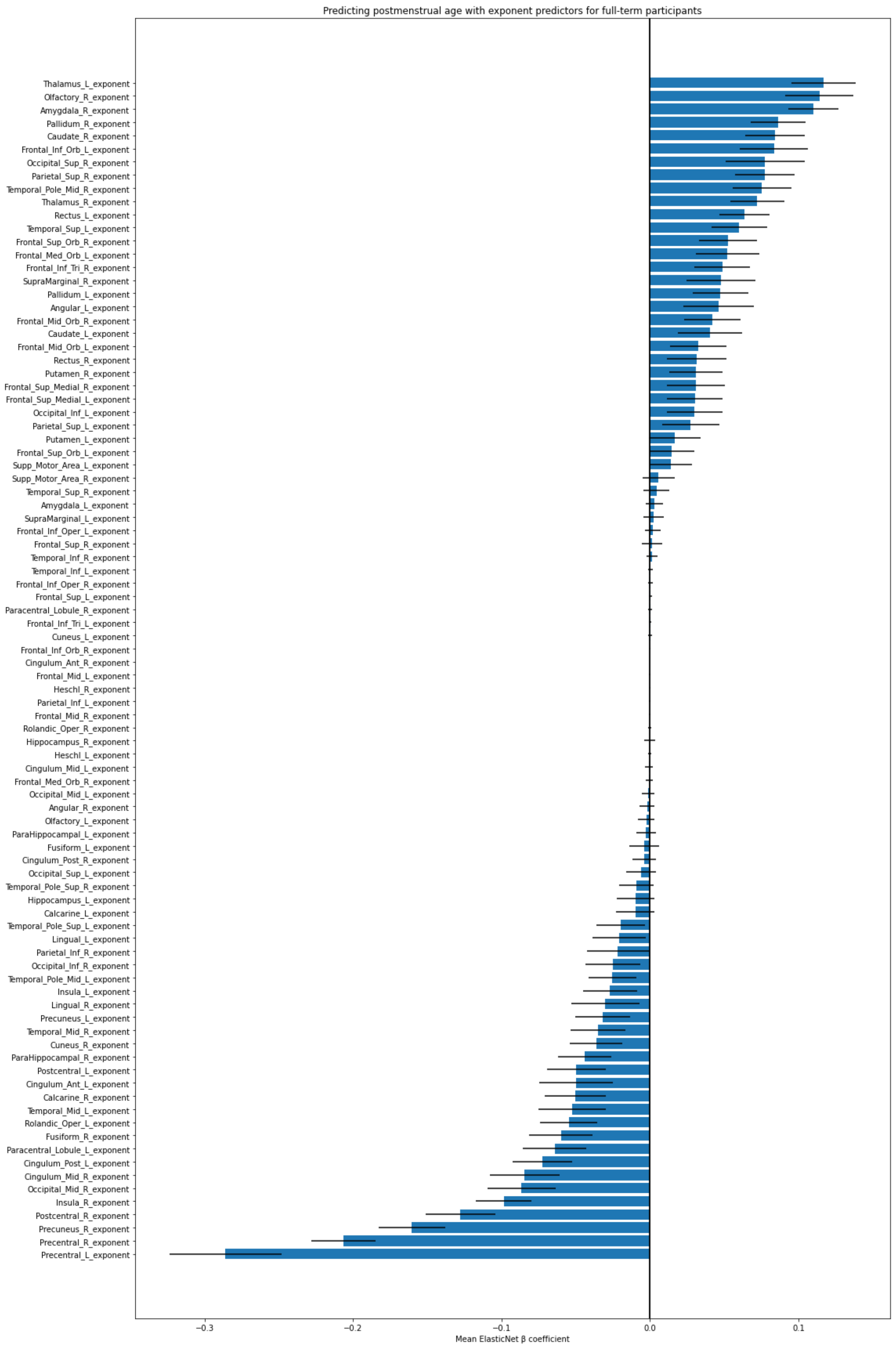


**Figure S11.** Exponent feature mean coefficients for predicting postmenstrual age at scan. Full-term subjects.
